# Increased circulating fibronectin, depletion of natural IgM and heightened EBV, HSV-1 reactivation in ME/CFS and long COVID

**DOI:** 10.1101/2023.06.23.23291827

**Authors:** Zheng Liu, Claudia Hollmann, Sharada Kalanidhi, Arnhild Grothey, Sam Keating, Irene Mena-Palomo, Stephanie Lamer, Andreas Schlosser, Agnes Kaiping, Carsten Scheller, Franzeska Sotzny, Anna Horn, Carolin Nürnberger, Vladimir Cejka, Boshra Afshar, Thomas Bahmer, Stefan Schreiber, Jörg Janne Vehreschild, Olga Miljukov, Christian Schäfer, Luzie Kretzler, Thomas Keil, Jens-Peter Reese, Felizitas A Eichner, Lena Schmidbauer, Peter U Heuschmann, Stefan Störk, Caroline Morbach, Gabriela Riemekasten, Niklas Beyersdorf, Carmen Scheibenbogen, Robert K Naviaux, Marshall Williams, Maria E Ariza, Bhupesh K Prusty

**Affiliations:** Institute for Virology and Immunobiology, Julius-Maximilians-University of Würzburg, Würzburg, Germany; Stanford Genome Technology Center, Stanford University School of Medicine, Stanford, CA, USA; Rudolf Virchow Center, Center for Translational Bioimaging, Julius-Maximilians-University of Würzburg, Germany; Institute for Medical Immunology, Charité-Universitätsmedizin Berlin, Berlin, Germany; Institute of Clinical Epidemiology and Biometry, Julius-Maximilians-University of Würzburg, Würzburg, Germany; Department of Clinical Research & Epidemiology, Comprehensive Heart Failure Center and Department of Medicine I, University Hospital Würzburg, Würzburg, Germany; Internal Medicine Department I, University Hospital Schleswig-Holstein UKSH - Campus Kiel, Kiel, Germany; University of Cologne, Faculty of Medicine and University Hospital Cologne, Department of Internal Medicine, Center for Integrated Oncology Aachen Bonn Cologne Duesseldorf, Germany; University Medicine Greifswald, Institute of Clinical Chemistry and Laboratory Medicine, Greifswald, Germany; Charité - Universitätsmedizin Berlin and Berlin Institute of Health (BIH), Berlin, Germany; Klinik für Rheumatologie, Universitätsklinikum Schleswig-Holstein, Lübeck; Departments of Medicine, Pediatrics, and Pathology, University of California, San Diego School of Medicine, San Diego, USA; Institute for Behavioral Medicine Research (IBMR), The Ohio State University, Columbus, Ohio, USA; Institute for Medical Data Sciences, University Hospital Würzburg, Würzburg; Clinical Trial Center, University Hospital Würzburg, Würzburg

**Keywords:** ME/CFS, long COVID, (n)IgM, Fibronectin, EBV, HSV-1, HHV-6, dUTPase

## Abstract

Myalgic Encephalomyelitis/ Chronic Fatigue syndrome (ME/CFS) is a complex, debilitating, long-term illness without a diagnostic biomarker. ME/CFS patients share overlapping symptoms with long COVID patients, an observation which has strengthened the infectious origin hypothesis of ME/CFS. However, the exact sequence of events leading to disease development is largely unknown for both clinical conditions. Here we show antibody response to herpesvirus dUTPases, particularly to that of Epstein-Barr virus (EBV) and HSV-1, increased circulating fibronectin (FN1) levels in serum and depletion of natural IgM against fibronectin ((n)IgM-FN1) are common factors for both severe ME/CFS and long COVID. We provide evidence for herpesvirus dUTPases-mediated alterations in host cell cytoskeleton, mitochondrial dysfunction and OXPHOS. Our data show altered active immune complexes, immunoglobulin-mediated mitochondrial fragmentation as well as adaptive IgM production in ME/CFS patients. Our findings provide mechanistic insight into both ME/CFS and long COVID development. Finding of increased circulating FN1 and depletion of (n)IgM-FN1 as a biomarker for the severity of both ME/CFS and long COVID has an immediate implication in diagnostics and development of treatment modalities.

## Main Text

Myalgic encephalomyelitis/chronic fatigue syndrome (ME/CFS) is considered as a chronic post viral illness that shares several overlapping clinical symptoms with long COVID or post-acute sequelae of SARS-CoV-2 infection (PASC)^1^ including neurological disturbances, extreme fatigue, post-exertional malaise (PEM), and postural orthostatic tachycardia syndrome (POTS). Immune dysregulation, microbiota dysbiosis, autoimmunity, vascular dysfunction, dysfunctional neurological signaling are some of the key hypothesized mechanisms for both the diseases^1^. However, there are no diagnostic biomarkers for the disease diagnosis and no treatment modalities yet. The infectious origin hypothesis of ME/CFS is long postulated and is supported by development of long COVID after SARS-CoV-2 infection. Our laboratories have postulated the idea of herpesvirus reactivation as a key mechanism of both ME/CFS as well as long COVID development, which is now corroborated by several groups^2, 3^. In this paper, we provide evidence for frequent HSV-1 and EBV reactivation in both ME/CFS and long COVID patients and provide an experimental reasoning for potential cellular damage through herpesvirus dUTPase proteins. Furthermore, we show that ME/CFS patients have altered autoimmune features, which possibly developed because of depletion of natural IgM ((n)IgM) within primary hematopoietic organs. Our study further focuses on fibronectin protein that shows altered expression and immune response patterns that correlates with disease severity.

## Results

### Increased HSV-1 and EBV reactivation in ME/CFS and long COVID

Reactivation of different herpesviruses including HHV-6, HHV-7 and EBV is frequently associated with ME/CFS development^4, 5^. Most recently, reactivation of several herpesviruses including EBV was detected during acute SARS-CoV-2 infection^2^. To test the possibility that these dormant viruses are being reactivated in long COVID patients and are producing dUTPases, we examined the humoral response against the viral dUTPases of EBV, HHV-6 and HSV-1 in a cohort of patients (n=278) at least after 6 months of first SARS-CoV-2 infection (COVIDOM cohort^6^). Only 6% of the patients of this cohort were hospitalized during the acute SARS-CoV-2 infection while rest of the 94% of the patients had mild to moderate SARS-CoV-2 infection without the need for hospitalization^7^. This patient cohort was further divided into three groups based on a scoring system^6^, which separated mild and severe long COVID patients (mild (n=107) and severe LC (n=22) respectively) from those who did not show any major health issues post SARS-CoV-2 infection and recovery (No LC, n=149). Distribution of these patient and control groups according to their age and gender is shown in Extended Fig. 1a (Supplementary Table 1). The long COVID cohort were compared to healthy blood donors (HC, n=31) and ME/CFS patients (ME/CFS, n=77). As commercial dUTPase ELISA based assay is not available, we used purified recombinant dUTPase proteins as a bait to capture human IgG in Western blots (Fig. 1a). Our results showed heightened IgG responses against EBV and HSV-1 dUTPases in ME/CFS and long COVID patients (Fig. 1b, Extended Fig. 1b). Strikingly higher IgG response to HSV-1 dUTPase was detected in mild and severe long COVID patients (Fig. 1b). A modest but significant increase in IgG response against HHV-6 dUTPase was observed in ME/CFS patients (Fig. 1b, Extended Fig. 1) but not in the cohorts of SARS-CoV-2 infected patients, which had decreased IgG response against HHV-6 dUTPase in comparison to healthy controls. These results suggest an overall increase in EBV and HSV-1 reactivation in both ME/CFS and SARS-CoV-2 infected patients.

**Fig. 1:**
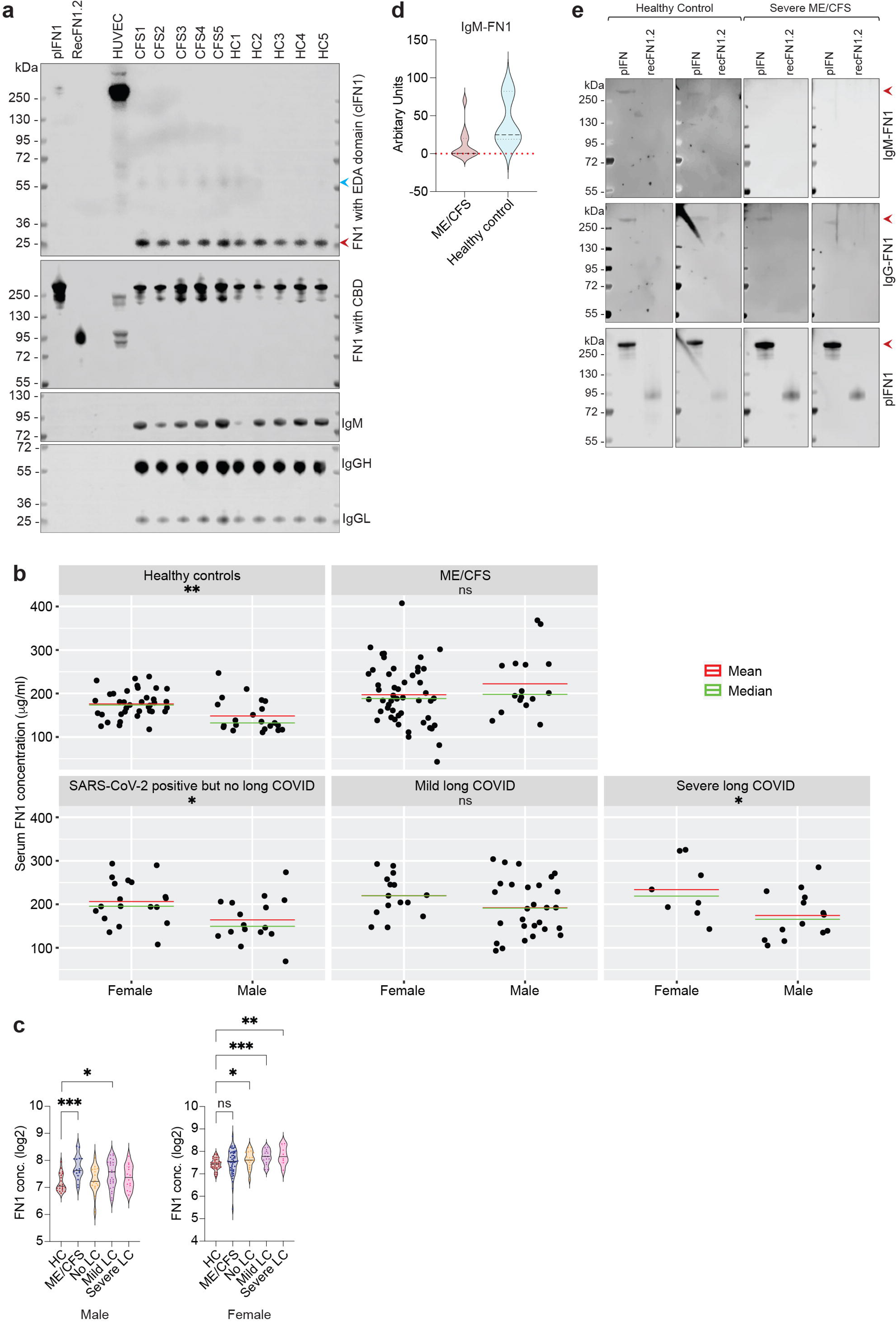
Reactivation of EBV in ME/CFS and long COVID patients and the role of viral dUTPase protein in cellular manipulation. **a**. Immunoblot based detection of IgG against herpesvirus dUTPase in human serum. Virus-specific recombinant protein bands detected by IgG are indicated. **b**. Likert chart showing percentage of positivity for antibodies against EBV, HSV-1 and HHV-6 dUTPase within healthy controls (HC), ME/CFS, Covid-19 PCR positive but without long COVID (No LC), mild LC and severe LC patients. Kruskall Wallis (and its higher order equivalent, ScheirerRayHare Test) rank sum test for antibody state. EBV, *P = 0.015; HSV-1, *P = 0.068; HHV-6, **P = 0.002. Mann-Whitney non-parametric test for EBV dUTPase HC vs ME/CFS, ***P = 0.0008; for HSV-1 dUTPase HC vs severe LC, P = 0.051; for HSV-1 dUTPase No LC vs severe LC, *P = 0.013. Different amounts IgG levels in patient serum was arbitrarily divided into 4 groups (0, absent; 1, low; 2, moderate; 3, high). **c.** Confocal images shows hyperpolarization of mitochondria in HEK293 cells under transient expression of HSV-1, HHV-6 and EBV dUTPases. Mock vector backbone was used as a control. **d.** Average mitochondrial surface area from 5 biological replicates are plotted in the form of a scatter plot. n=5. Unpaired two-tailed non-parametric t-test. Mock vs EBV dUTPase, *P = 0.0414; Mock vs HHV-6 dUTPase, **P = 0.0077; Mock vs HSV-1 dUTPase, *P = 0.0479. **e.** Immunoblot analysis shows increase in mitofusin1 (Mfn1) and decrease in LC3β protein levels in presence of herpesvirus dUTPases. GAPDH staining was used as a loading control. Mean Mfn1 and LC3β protein levels are presented as scatter plots. Data from 3 independent experiments. n=3. Unpaired two-tailed non-parametric t-test. For Mfn1, *P = 0.03 (Mock vs HHV-6); *P = 0.01 (Mock vs HSV-1). For LC3β, *P = 0.01 (Mock vs EBV); *P = 0.05 (Mock vs HHV-6). **f.** EBV dUTPase interferes with autophagosome assembly. **g.** TMRE dyes were used to study mitochondrial membrane potential and OXPHOS in HEK293 cells. Cells were transiently transfected with herpesvirus dUTPases or a mock vector for 48 h. Trypsinized cells were stained with TMRE dye and were used for flow cytometry. Oligomycin was used to inhibit ATP synthase. Data from 3 independent experiments. n=3. MFI, mean fluorescence intensity. **h.** Normalized log2 ratio of LFQ (label-free quantitation) intensities of proteins. Fold change of proteins in HSV-1 vs Mock was plotted against the same in EBV vs Mock to highlight proteins that were common and were enriched in both sample sets. Circles indicate identified cellular proteins; circle size correlates with the number of razor and unique peptides used for quantification. Significantly enriched proteins that are potential interaction partners of EBV and HSV-1 dUTPases are displayed in red. **i.** Immunoblot analysis to validate potential herpesvirus dUTPase interacting partners identified from co-IP. GAPDH staining was used as a negative control.

### Herpesvirus dUTPase proteins alters mitochondrial architecture

The increased anti-dUTPase IgG response observed in patient cohorts after 6 months of first SARS-CoV-2 infection suggests EBV and HSV-1 reactivation potentially contributing to disease development. Studies by our group have shown that the herpesviruses dUTPases represent a new family of Pathogen Associated Molecular Patterns (PAMPs)^7^ protein ligands with novel immune and neuromodulatory properties, independent of the enzymatic activity. Hence, we further explored the potential role of these viral proteins in human biology.

Transient expression of HSV-1, HHV-6 and EBV dUTPase in cultured human cells induced a hyperpolarized and hyperfused mitochondrial phenotype (Fig. 1c, Extended Fig. 2a) with increased mitochondrial surface area (Fig. 1d, Extended Fig. 2b). The appearance was similar to the stress-induced mitochondrial hyperfusion (SIMH) phenotype that is seen with exposure to certain environmental chemicals^8^, nutrient depletion, viral infections, and in certain chronic disease states^9^. In U2-OS cells, all the three dUTPases were predominantly localized within the nucleus (Extended Fig. 2a). In HEK293 cells, only EBV dUTPase localized to the nucleus whereas HSV-1 and HHV-6 dUTPases remained mostly within cytoplasm (Fig. 1c). EBV dUTPase (BLLF3) is known to be localized within nucleus^10^ whereas HSV-1 dUTPase (UL50) is reported to be localized within both cytoplasm and nucleus. Irrespective of the protein localization, mitochondrial fusion protein Mfn1 was upregulated by all the three viral dUTPases (Fig. 1e). Other mitochondrial fusion proteins like Mfn2, Miga1, remained unchanged (Extended Fig. 2c). Mitophagy was inhibited to various degree by all the three dUTPases, EBV dUTPase being the most efficient in decreasing LC3β protein expression (Fig. 1e). Tetramethylrhodamine ethyl ester (TMRE) staining of dUTPase transfected cells in the presence of ATP synthase inhibitor, oligomycin showed a decrease in mitochondrial membrane potential in cells expressing EBV dUTPase (Fig. 1f) suggesting poor mitochondrial health and energetics particularly in presence of EBV dUTPase. HSV-1 and HHV-6 dUTPase did not show any significant effect (Extended Fig. 2d). EBV dUTPase expressing cells showed characteristics of hyperpolarized mitochondria with higher mitochondrial membrane potential in comparison to mock vector transfected cells in the absence of oligomycin (Extended Fig. 2e). On the other hand, exposing U2-OS cells in culture to recombinant EBV dUTPase induced Drp-1 dependent mitochondrial fragmentation and appearance of spherical hyperpolarized mitochondria (Extended Fig. 2f-g) in a dose dependent manner that showed increased mitochondrial membrane potential under normal growth conditions, which decreased in presence of oligomycin suggesting poor mitochondrial health (Extended Fig. 2h). Such spherical mitochondrial phenotype was previously described as ‘mitosphere’ that induces oxidative stress^11^. We constitutively expressed all the three viral dUTPases and immunoprecipitated viral dUTPase proteins in HEK293 cells (Extended Fig. 2i). Mass spectrometry analysis of co-immunoprecipitated proteins showed enrichment of several cellular cytoskeleton associated proteins including neurofilament medium chain (NEFM), epiplakin 1 (EPPK1), plectin (PLEC) (Fig. 1g-h, Extended Fig. 2j-k) suggesting strong association of host cell cytoskeleton with herpesvirus dUTPases. Cellular localization of dUTPase proteins seems to be not associated with its interaction with cell cytoskeleton as both HSV-1 dUTPase and EBV dUTPase showed similar interacting partners. We further validated some of the interacting partners of EBV and HSV-1 dUTPases identified by mass spectrometry through immunoblotting (Fig. 1h, Extended Fig. 2l). These results suggest that herpesvirus dUTPases interact with host cell cytoskeleton and alter mitochondrial architecture as well as function.

### Immunoglobulins and mitochondrial fragmentation in ME/CFS

Chronic, recurrent HSV-1 and EBV infections lead to development of autoimmunity^12,13^. We hypothesized that autoimmunity-induced antibodies in serum might induce mitochondrial dysfunction as shown by others^14, 15^. To check if immunoglobulins (Igs) isolated from ME/CFS patients can induce mitochondrial alterations *in vitro*, we purified Ig fractions from ME/CFS patients (n=17) and healthy controls (n=13) using Sepharose G columns. Exposure of as low as 1 μg of purified immunoglobulins from severe ME/CFS patients fragmented mitochondria within 12 h of exposure (Fig. 2a-b). The mitochondrial fragmentation phenotype for severe ME/CFS patients was cell-type dependent and was pronounced in primary human umbilical vein endothelial cells (HUVEC) (Fig. 2a-b). Ig from healthy controls showed a mixed effect on mitochondrial architecture whereas Ig from mild/moderate ME/CFS patients had a modest but significant effect on mitochondria in HUVEC cells. Fragmented mitochondrial phenotype by Ig from severe ME/CFS patients showed decreased Mfn1 and PLD6 protein levels (Fig. 2c). Efficiency of IgG purification was tested by analyzing antibodies against EBV dUTPase in EBV positive and negative patient serum, which showed enriched EBV dUTPase antibodies after purification (Extended Fig. 3a, b).

**Fig. 2:**
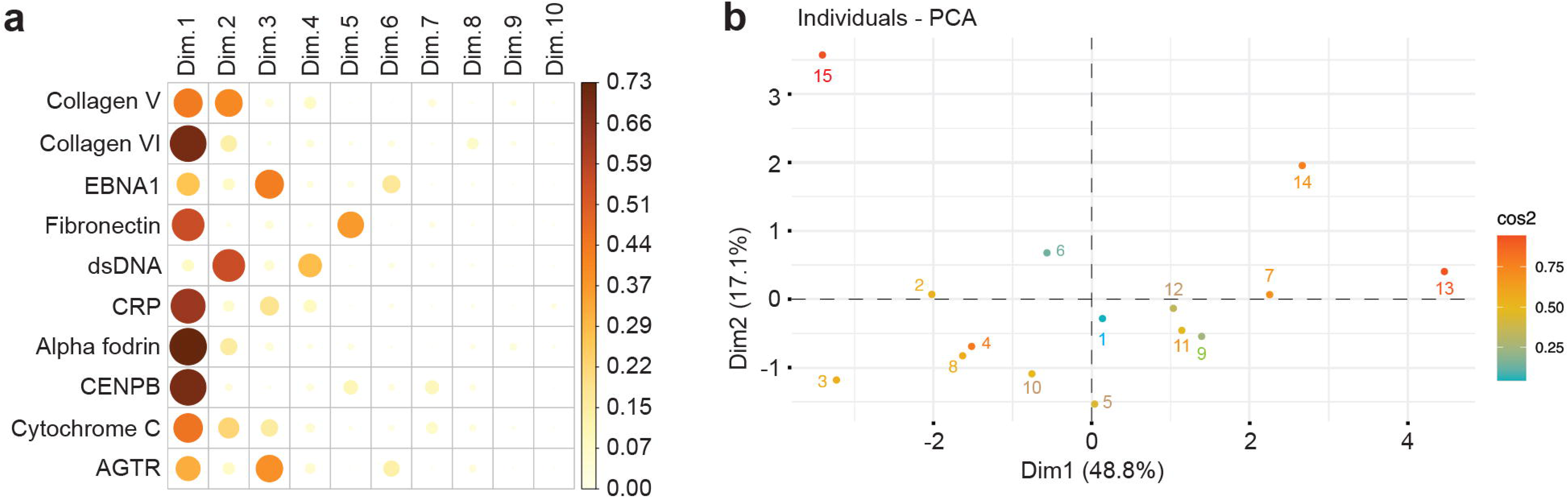
Autoimmunity, mitochondrial alterations and circulating Fibronectin levels in ME/CFS. **a.** Confocal images show mitochondrial architecture in primary HUVEC cells exposed to 1μg of purified IgG from patient sera. Two different representative images for each condition is shown. **b.** Average mitochondrial surface area in primary HUVEC cells exposed to 1μg of purified IgG from patient sera is quantified and compared between healthy controls, mild/moderate, severe ME/CFS. Data from three independent experiments from each serum sample is shown as a violin plot. n=3. Two-tailed non-parametric test. Healthy control vs mild/moderate ME/CFS, *P = 0.0329. Healthy control vs severe ME/CFS, **P = 0.0046. Mild/moderate vs severe ME/CFS, ****P < 0.0001. **c.** Immunoblot analysis shows decrease in mitofusin1 (Mfn1) and PLD6 protein levels in HUVEC cells exposed to 1μg of purified IgG from patient sera for 12 h. Actin staining was used as a loading control. Fold change values were derived from densitometric analysis of bands after normalization with the same for actin. n=2. HD, healthy donors; CFS, severe CFS patients. **d**. Heat map of log2 fold LFQ intensities of proteins detected within purified immune complexes from patient sera. Three proteins that showed differential protein levels between healthy controls and ME/CFS patients are shown. **e**. Multivariate analysis of clusters based on distance metrics derived from IgM antibody levels for a panel of autoantigens. Log-transformed scaled data showing relative differences between different variable in both healthy controls and patients. **f**. The Variables Factor map for the Principal Components (for Patients and Healthy Controls combined data) shows the projection of the top 10 Autoantigen variables projected onto the plane spanned by the first two Principal Components. **g**. Serum fibronectin (FN1) levels in patient sera. Log2 values of FN1 are presented as a violin plot. Two-tailed parametric t-test. Healthy control (HC) vs ME/CFS, **P = 0.005. **h.** Kernel density plot showing the bivariate serum FN1 distributions among healthy controls and ME/CFS patients. FN1 concentrations on both X- and Y-axis are presented as μg/ml. **i.** Circulating fibronectin (FN1) levels correlates with ME/CFS severity associated Bell score. Log2 fold FN1 vales are presented as a violin plot. Two-tailed parametric t-test. HC vs Bell 0-20, ****P < 0.0001. Bell 0-20 vs Bell 30-50, **P = 0.0032. **j.** AUROC analysis for circulating FN1 levels in healthy controls (HC) vs severe ME/CFS patients. **k.** Serum fibronectin (FN1) levels in different patient groups post SARS-CoV-2 infection. Log2 values of FN1 are presented as a violin plot. Two-tailed parametric t-test. Healthy control (HC) vs mild LC, **P = 0.0032. HC vs severe LC, *P = 0.0488. ns, not significant.

Purified IgG from human serum mostly contains immunoglobulin fractions with freely available Fc receptors along with associated immune complex proteins. These immune complex proteins can also potentially be a factor that can induce mitochondrial fragmentation. Hence, we analyzed the same purified IgG fractions from healthy control (n=12) and ME/CFS patients (n=15), that were used in mitochondrial studies, by mass spectrometry. IgG and IgM bound immune complex purification was of high quality and uniform throughout the samples as validated by similar enrichment of complement components in all the samples (Extended Fig. 3c). Only three cellular proteins showed decreased amounts within the immune complex of ME/CFS patient groups in comparison to healthy controls (Fig. 2d, Extended Fig. 3d), i.e, fibronectin (FN1), alpha2 macroglobulin (A2M) and serotransferrin (TF).

In an independent microarray study, we also detected IgM against fibronectin antigen as one of the few IgMs that were selectively decreased in severe ME/CFS patients. To test potential IgM response against selective autoantigens frequently involved in autoimmune diseases, IgM levels were tested against 120 autoantigens (Supplementary Table 2) in a small cohort of mild to severe ME/CFS patients (n=12) and healthy controls (n=3). Samples were blinded throughout the experimental procedure and data analysis. Multivariate clustering of log-transformed data resulted in the observation that patients could be separated into distinct groups comprising healthy, mild/moderate and severe patients (Fig. 2e) on the basis of IgM antibody levels against autoantigens like PCNA, collagen V and VI, complement C3, CRP etc. (Fig. 2f, Extended Fig. 4a-b). IgM against fibronectin was one of the 10 variables that contributed to these clustering. These results suggest that decreased immunoglobulin against fibronectin and other proteins are key features of severe ME/CFS.

### ME/CFS and long COVID patients show increased amounts of circulating fibronectin

FN1 interacts with circulating immune complex and aggregates with IgG, IgM and IgA thereby playing a significant role in both the clearance of activated immune complexes and tissue deposition of fibronectin-containing immune complexes in several diseases^16^. Lack of fibronectin within immune complexes and their potential accumulation within blood can indicate lack of protection against certain pathogenic infections. To check if overall fibronectin protein expression is downregulated in ME/CFS or is specifically removed from immune complexes for some reason, we measured circulating fibronectin levels in serum of ME/CFS (n=66) patients and healthy controls (n=63). Circulating fibronectin levels were significantly higher in ME/CFS patients in comparison to healthy controls as shown in the Kernel Density plot (Fig. 2g-h). Serum FN1 levels showed a positive correlation with ME/CFS severity as patients with bell scores of 0-20 had significantly higher FN1 levels (Fig. 2i, HC vs Bell 0-20, ****P < 0.0001. Bell 0-20 vs Bell 30-50, **P = 0.0032) in serum than those with bell score between 30-50 (Area under the receiver operator characteristic (AUROC) = 79.8% for severe ME/CFS (Fig. 2j), P<0.0001). We further compared circulating fibronectin levels within SARS-CoV-2 positive cohorts. Only long COVID patients (both mild and severe) showed significantly higher circulating fibronectin levels (Fig. 2k, HC vs mild LC, **P = 0.0032. HC vs severe LC, *P = 0.0488) in comparison to healthy controls and SARS-CoV-2 positive but without long COVID individuals (No LC). Circulating fibronectin contains the majority of plasma fibronectin (plFN1) secreted by hepatocytes and a minor fraction of cellular fibronectin (clFN1) secreted by other cell types. We compared circulating FN1 composition between some of the ME/CFS patients to that of healthy controls, which showed an overall increase in both extra domain A (EDA) negative plFN1 as well as EDA domain and cell binding domain (CBD) positive clFN1 in ME/CFS patients (Extended Fig. 5a) suggesting an overall increase in circulating fibronectin levels. Total human IgG was used as loading control. Interestingly, we observed significant differences in circulating fibronectin levels between male and females (Extended Fig. 5b). Particularly, healthy controls and SARS-CoV-2 positive but no long COVID (no LC) groups showed significantly higher FN1 levels in females than males (Extended Fig. 5b, HC male vs female, **P = 0.0014; No LC male vs female, *P = 0.0204; severe LC male vs female, *P = 0.0347). Similarly, increasing FN1 levels among female long COVID patients showed positive correlation with disease severity (Extended Fig. 5c, HC male vs ME/CFS male, ***P = 0.0001; HC male vs mild LC male, *P = 0.012; HC female vs no LC female, *P = 0.0193; HC female vs mild LC female, ***P = 0.0006; HC female vs severe LC female, **P = 0.0016), which was less among male long COVID patients. However, because of the comparatively low levels of circulating FN1 among healthy males, male ME/CFS patients showed a significant increase in FN1 levels (Extended Fig. 5c). In summary, our results show increased circulating FN1 levels in ME/CFS and long COVID patients that correlates with disease severity.

### Depletion of IgM against FN1 correlates with disease severity

One of the interesting observations from our antibody microarray studies was the decrease in IgM antibodies against FN1 among some of the severe ME/CFS patients (Fig. 2a, Extended Fig. 5d). Antibodies (IgG, IgM and IgA) against FN1 are frequently detected in plasma of healthy individuals^17^. Moreover, all the three forms of immunoglobulins can bind to the same region of the FN1 protein^16^. An inverse correlation between increasing circulating FN1 levels and decreased IgM responses against FN1 were previously documented during Trypanosoma infection^18^. Hence, we argued that IgM against FN1 possibly belongs to the natural IgM ((n)IgM) category. Natural IgM are key immunoglobulins produced specifically by plasma B1 B cells^19^ and have important scavenger and protector function^20^. Decreased (n)IgM can be the source for autoimmunity^19^. As a proof of concept, we validated the microarray results using immunoblotting with purified human plFN1 and recombinant FN1.2 proteins as a bait, in 7 severe ME/CFS, 5 long COVID and 5 healthy controls, which confirmed the microarray data showing undetectable or poorly detectable amounts of IgM-FN1 (Extended Fig. 5e) in severe ME/CFS and long COVID patients. IgM-FN1 was detectable in healthy controls. It is noteworthy that human IgM detected only the full-length plFN1. For a quantitative analysis, we further developed sandwich ELISA assays to measure serum IgM and IgG levels against human FN1 and measured both in healthy controls (n=63), ME/CFS (n=66) and three different groups of SARS-CoV-2 positive patients (no LC, n=55; mild LC, n=63; severe LC, n=22). The total IgM-FN1 levels were not different between healthy controls and ME/CFS patients (Fig. 3a). However, SARS CoV-2 positive patients showed significantly decreased IgM-FN1 levels in comparison to both healthy controls and ME/CFS patients (Fig. 3a, HC vs No LC, HC vs mild LC, HC vs severe LC, ****P < 0.00001. No LC vs severe LC, *P = 0.0376). Furthermore, IgM-FN1 amounts correlated with long COVID severity with the strongest decrease in severe LC patients (Fig. 3a). Similarly, IgG against FN1 was also significantly decreased in all the three groups of SARS-CoV-2 positive patients (Fig. 3b, HC vs No LC, HC vs mild LC, HC vs severe LC, ****P < 0.00001). Severe LC patients showed a trend towards recovery of IgG-FN1 levels. Further separation of ME/CFS patients based on disease severity (Bell score), showed significantly decreased IgM-FN1 levels only in severe patients with a Bell score between 0-20 (Fig. 3c, HC vs Bell 0-20, **P = 0.0046. Bell 0-20 vs Bell 30-50, ***P = 0.0002). IgG-FN1 did not show any association with ME/CFS disease severity (Fig. 3d). To answer whether specifically (n)IgM against fibronectin was depleted post SARS-CoV-2 infection or not, we tested two of the most common (n)IgMs against phosphoryl choline (PC) and Malondialdehyde (MDA) in a small cohort of healthy controls and all the three groups of SARS-CoV-2 positive patients (n=20 for each group). Both IgM-PC and IgM-MDA levels showed a significant correlation with long COVID severity (Fig. 3e-f, Extended Fig. 6a-b) being the lowest in severe LC patients. AUROC analysis of IgM-FN1 also showed increased detection accuracy as the severity of long COVID increased from no LC to severe LC (Fig. 3g-i). Muti-variate ROC analysis using both circulating FN1 and IgM-FN1 levels showed that both these proteins together can potentially serve as a biomarker for severe ME/CFS (Fig. 4a, accuracy of 83.7%) and severe long COVID (Fig. 4b, accuracy of 85.3%). Multiple logistic regression analysis separated healthy controls from severe ME/CFS patients based on both FN1 and IgM-FN1 levels with 83.7% accuracy (Fig. 4c) and healthy controls from both mild and severe long COVID patients with 83.3% accuracy (Fig. 4d). Moreover, both ME/CFS and long COVID patients showed close similarity in terms of their FN1 and IgM-FN1 amounts (Fig. 4e). In fact, severe groups of both ME/CFS and long COVID patients closely resembled each other (Fig. 4f). Interestingly, all the three natural IgMs, i.e., (n)IgM-FN1, (n)IgM-PC and (n)IgM-MDA levels showed a trend towards being more abundant in healthy females than males (Extended Fig. 7a-b). In fact, female ME/CFS patients showed a significant decrease in IgM-FN1 than male patients (Extended Fig. 7a) which was insignificant among other patient groups. All the natural IgM levels were significantly lower in all three groups of LC patients (Extended Fig. 7c) in comparison to healthy controls and had no clear association with gender. In summary, our results show strong depletion of (n)IgMs following SARS-CoV-2 infection even after 6 months post-virus infection. Significantly increased circulating fibronectin levels together with the decreased (n)IgM response against fibronectin has the potential to separate SARS-CoV-2 infected non-long COVID individuals from long COVID and severe ME/CFS patients.

**Fig. 3:**
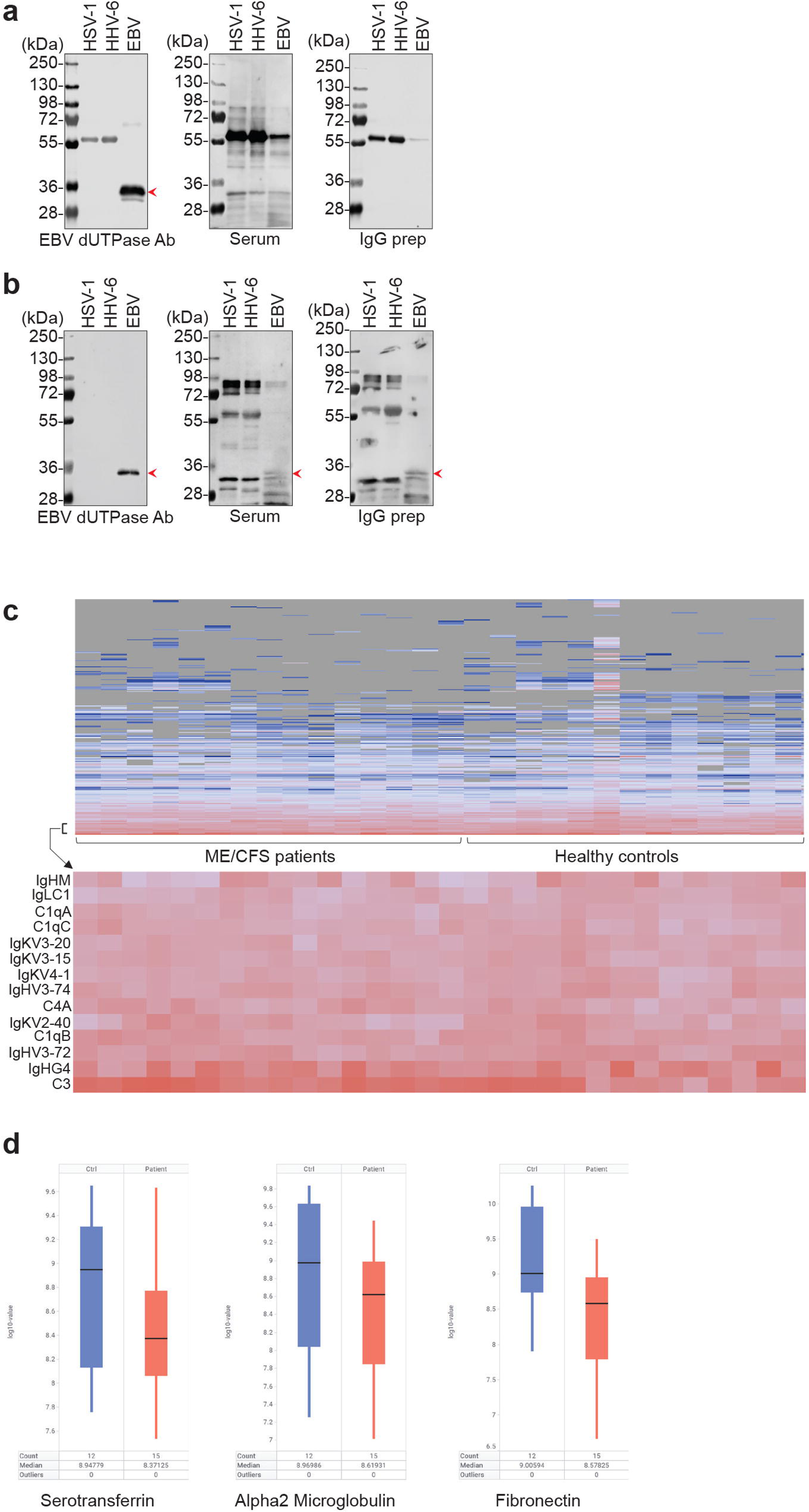
IgM and IgG levels against Fibronectin in ME/CFS and long COVID. **a.** IgM levels against fibronectin (IgM-FN1) in patient sera. Log2 fold IgM-FN1 amounts are presented as a violin plot. Two-tailed parametric t-test. Healthy control (HC) vs SARS CoV-2 positive but without long COVID (No LC), HC vs mild LC, HC vs severe LC, ****P < 0.00001. No LC vs severe LC, *P = 0.0376. ns, not significant. **b.** IgG levels against fibronectin (IgM-FN1) in patient sera. Log2 fold IgG-FN1 amounts are presented as a violin plot. Two-tailed parametric t-test. Healthy control (HC) vs SARS CoV-2 positive but without long COVID (No LC), HC vs mild LC, HC vs severe LC, ****P < 0.00001. ns, not significant. **c.** IgM-fibronectin (FN1) levels correlates with ME/CFS severity associated Bell scores. Log2 fold IgM-FN1 vales are presented as a violin plot. Two-tailed parametric t-test. HC vs Bell 0-20, **P = 0.0046. Bell 0-20 vs Bell 30-50, ***P = 0.0002. **d.** IgG-fibronectin (FN1) levels does not correlate with ME/CFS severity associated Bell scores. Log2 fold IgG-FN1 vales are presented as a violin plot. Two-tailed parametric t-test. Ns, not significant. **e.** IgM levels against phosphorylcholine (IgM-PC) in patient sera. Log2 fold IgM-PC amounts are presented as a violin plot. Two-tailed parametric t-test. Healthy control (HC) vs mild LC, ***P = 0.0002. HC vs severe LC, ***P = 0.0006. No LC vs severe LC, *P = 0.0216. No LC vs mild LC, *P = 0.0111. ns, not significant.\ **f.** IgM levels against malondialdehyde (IgM-MDA) in patient sera. Log2 fold IgM-MDA amounts are presented as a violin plot. Two-tailed parametric t-test. Healthy control (HC) vs mild LC, *P = 0.0175. HC vs severe LC, **P = 0.0068. No LC vs severe LC, *P = 0.0209. No LC vs mild LC, *P = 0.0525. ns, not significant. **g.** AUROC analysis for IgM-FN1 levels in healthy controls (HC) vs SARS CoV-2 positive but without long COVID (No LC) patients. **h.** AUROC analysis for IgM-FN1 levels in healthy controls (HC) vs mild LC patients. **i.** AUROC analysis for IgM-FN1 levels in healthy controls (HC) vs severe LC patients.

**Fig. 4:**
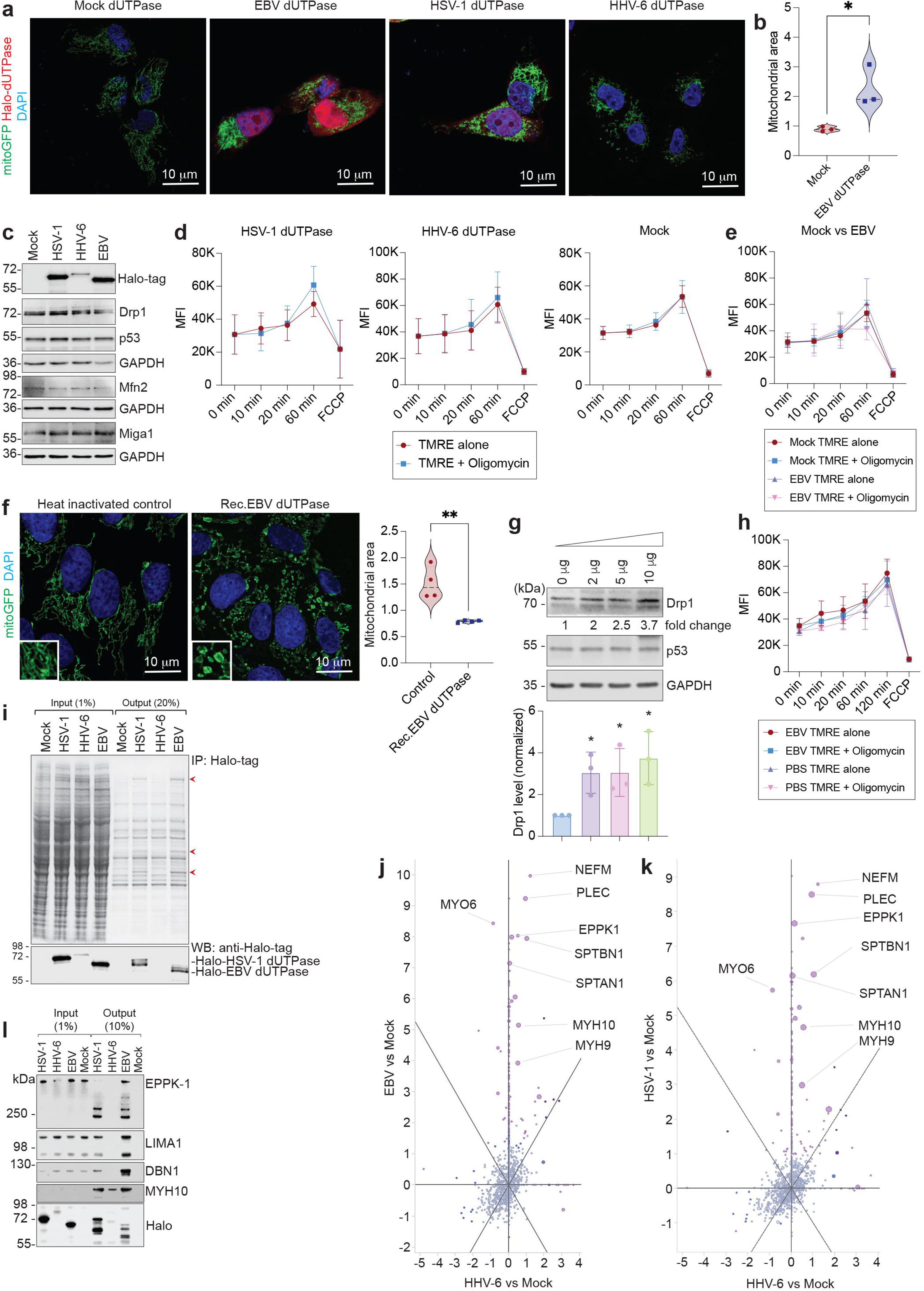
IgM-FN1 and circulating fibronectin as a biomarker in ME/CFS and long COVID. **a.** Multi-variate AUROC analysis for circulating FN1 and IgM against fibronectin (IgM-FN1) in healthy controls (HC) vs severe ME/CFS patients. Multiple logistic regression. ****P < 0.0001. **b.** Multi-variate AUROC analysis for circulating FN1 and IgM against fibronectin (IgM-FN1) in healthy controls (HC) vs severe LC patients. Multiple logistic regression. ****P < 0.0001. **c.** Multiple variable bubble plot with circulating FN1 levels plotted against IgM-FN1 for healthy controls (HC) and severe ME/CFS patients. **d.** Multiple variable bubble plot with circulating FN1 levels plotted against IgM-FN1 for healthy controls (HC) and total long COVID patients (mild LC + severe LC). **e.** Multiple variable bubble plot with circulating FN1 levels plotted against IgM-FN1 for ME/CFS and total long COVID patients (mild LC + severe LC). **f.** Multiple variable bubble plot with circulating FN1 levels plotted against IgM-FN1 for severe ME/CFS (with Bell 0-20) and severe long COVID patients.

## Discussion

Chronic post viral illnesses like ME/CFS and long COVID affect multiple body systems and may lead to development of overlapping clinical features with other known health conditions making the diagnosis and treatment difficult for clinicians. No biomarker has been identified for either of the conditions to date^1^. Our study found increased humoral response against HSV-1, HHV-6 and EBV dUTPases in ME/CFS patients and the same against HSV-1 and EBV in long COVID patients suggesting herpesvirus reactivation as a potential contributor to both the clinical conditions. These results support our previous studies in ME/CFS patients^21^ and corroborate findings from other labs^3^ showing frequent EBV reactivation post COVID-19 infection. Chronic herpesvirus infection contributes to autoimmunity^12, 13^. Heightened expression of EBV dUTPase protein in plasma cell aggregates near glomeruli was shown in kidney biopsy tissue from lupus nephritis (LN) patients (class III and IV) undergoing flares that also exhibit an enrichment of long-lived autoantibody producing memory plasma cells in the kidneys^22^, which are believed to contribute to disease pathogenesis. Interestingly, the terminal differentiation of memory B-cells into plasma cells causes reactivation of latent EBV resulting in the expression of the dUTPase^23, 24^. The TLR2/MyD88/miRNA155/Ets-1 pathway is required to produce autoantibodies that form DNA-containing immune complexes (IgG anti-ssDNA, IgG anti-dsDNA)^25^ and EBV dUTPase is a potent activator of TLR2/MyD88 signaling and inducer of pro-inflammatory cytokines and miR155. In some ME/CFS patients exhibiting a dysfunctional GC Ab response, EBV dUTPase may contribute to this process by stimulating an extrafollicular Ab response that promotes increased splenic iNKTFH and marginal zone B cells, which could result in the formation of autoreactive B cells and, subsequently, the production of autoreactive Abs^26^. Interestingly, the dUTPase protein is also released in exosomes from EBV-infected B cells (plasma cells) during abortive lytic replication of EBV and these dUTPase-containing exosomes induced the production of pro-inflammatory cytokines in dendritic cells and PBMCs by engaging TLR2^27^. We argue that herpesvirus dUTPases post virus reactivation can play a contributing role in disease development. We have recently shown presence of EBV dUTPase proteins within brain biopsies of ME/CFS patients^5^ supporting the notion that strong neurological symptoms of ME/CFS potentially can be due to herpesviruses like HHV-6, EBV reactivating within specific tissues^28^. However, nearly uniform levels of depletion of IgM-FN1 and IgG-FN1 in all the SARS-CoV-2 positive patients studied suggest that herpesvirus reactivation alone can’t be responsible for the disease development. At the same time, a decrease in the IgG response against HHV-6 dUTPase in long COVID patients (Fig. 1b) could suggest alterations in the B cell response against specific pathogenic antigens post SARS-CoV-2 infection. We argue that ME/CFS and long COVID are post viral illnesses where the origin of the disease and acute post-infectious clinical features are potentially different. But both the diseases seem to overlap with each other sharing similar secondary chronic clinical features (Extended Fig. 8).

Endothelial cell dysfunction is a key clinical feature of both ME/CFS^29^ and long COVID^30^. In this study, immunoglobulins isolated from severe ME/CFS patients induced alterations in mitochondrial morphology of cultured primary cells favoring a fragmented mitochondrial phenotype. Further proteomic characterization of isolated immunoglobulins and associated circulating immune complexes (CIC) revealed decreased presence of three proteins within the IgG-bound CIC (namely TF, A2M and FN1), FN1 levels showing the clearest differences to that from healthy individuals. On the other hand, circulating FN1 was significantly increased in serum of ME/CFS patients. Atherosclerosis associated inflammation shows deposition of fibronectin in endothelial cells^31^. Antigens that bind to mesangial fibronectin induce glomerulonephritis through immune complex formations^32^. Plasma fibronectin is targeted by several bacteria including *Borrellia burgdorferi*^33^ and *S. aureus*^34^ for cell colonization. On the other hand, antibody response develops against plasma fibronectin specifically bound to early Borrelia antigen RevA^35^. Fibronectin is an essential component of circulating immune complexes^36^ because of its interaction with complement proteins like C1q^37^ and C3 that protect against pathogenic infections^38^. Hence, our finding of decreased fibronectin within immune complexes of ME/CFS patients could suggest poor bacterial and other pathogen clearance. Significantly increased circulating fibronectin in ME/CFS and long COVID patients was positively correlated with disease severity. Circulating fibronectin binds to TLR4 and induces an inflammatory cytokine response^39^. Specific domains of fibronectin also activate platelets^40^ and mast cells^41^. Increased serum/plasma fibronectin is commonly detected in rheumatic diseases^36^ and is suggested to be the cause of polyclonal activation of B cells^42^. Interestingly, irrespective of disease groups, female patients showed significantly increased circulating FN1 protein levels compared to male patients (Extended Fig. 5b) suggesting a possibility to reach the pathological threshold sooner than males. Increased circulating fibronectin levels are correlated with low levels of IgM against fibronectin during Trypanosoma infection^18^. Using different experimental approaches, we showed depletion of IgM against fibronectin as a characteristic feature of severe ME/CFS and long COVID suggesting depletion of IgM-FN1 as a key contributor to disease severity.

Natural IgM are critical players in maintaining cellular homeostasis through their scavenger, protector, and regulator function^20, 43^. Protection against cellular debris that can cause development of autoimmunity is a key function of (n)IgM. We argue that IgM against fibronectin (IgM-FN1) belongs to the (n)IgM category as IgM-FN1 is frequently detected in plasma of healthy individuals^17^. Our results showed presence of abundant IgM against FN1 in serum of healthy individuals supporting the notion that IgM-FN1 might belong to the natural IgM category and is crucial in complement activation and protection against autoimmunity. We showed that (n)IgM-FN1 binds only to full-length plasma fibronectin and does not have specificity for cellular fibronectin. As we found decreased fibronectin within the immune complexes of ME/CFS patients, we argue that severe ME/CFS and possibly long COVID patients have compromised complement activation against fibronectin leading to poor pathogen clearance and poor scavenging abilities against apoptotic cells causing generation of a broad range of autoimmune antibodies. Additionally, we noticed a decrease in two of the most common (n)IgMs against PC and MDA in SARS-CoV-2 positive patient cohorts, which strengthens the idea that loss of natural IgMs might be the cause of autoimmunity in both ME/CFS and long COVID patients. Additionally, we observed that females tend to have increased (n)IgM levels than males supporting the idea of immune differences between both genders^44^. These differences in circulating FN1 levels among male and female patients as well as gender-dependent differences in (n)IgM levels might explain the predominance of both ME/CFS and long COVID among females compared to males^1^.

Natural antibodies (IgM, IgG and IgA) are produced by innate-like B cells like B1, B2, and marginal zone (MZ) B cells^45^. (n)IgMs are specifically produced by B1 B cells within bone marrow and other primary lymphoid tissues^19^ and by circulating CD20^+^CD38^hi^ B cells^46^. We argue that under specific yet to be understood conditions, infection-mediated alterations within primary hematopoietic organs might lead to alterations in B1 B cells (Extended Fig. 8), which can eventually lead to depletion of natural IgM and development of autoimmunity. Measles virus also depletes memory B cells by an unknown mechanism leading to ‘immune amnesia’^47^. Another potential reason for depletion of (n)IgM can be alterations in CD4^+^ T cells. Non-cognate CD4^+^ T cells, which help plasma B1 cells in mice to generate a unique repertoire of (n)IgM^48^. Furthermore, CXCR4+ regulatory CD4^+^ T (Treg) cells manipulate both serum IgM levels and (n)IgM production by B1 B cells in the bone marrow^49^. T cell exhaustion and alterations in Treg numbers are documented in ME/CFS^50^. Hence modulation of T cell biology within primary hematopoietic tissues can have a potential impact on plasma B1 B cells and (n)IgM levels.

In summary, our findings reveal depletion of (n)IgM to be associated with disease severity in ME/CFS and long COVID, which may point to an autoimmune mechanism. Autoimmunity is considered as a key causal factor for diseases like ME/CFS. Hence first therapeutic interventions have been targeted to deplete pathogenic autoantibodies or B cells. In this regard our findings have immediate implications in both ME/CFS and long COVID diagnosis and further development of autoantibody treatment.

## Data Availability

Complete experimental data together with statistical data are uploaded to Mendeley (doi: 10.17632/4xkft5g9r5.1) and is currently under embargo till 13th June 2024. The data will be freely available to all the readers afterwards.

## Acknowledgements

The study was carried out using the clinical-scientific infrastructure of NAPKON (Nationales Pandemie Kohorten Netz, German National Pandemic Cohort Network) of the Network University Medicine (NUM), funded by the Federal Ministry of Education and Research (BMBF).

We gratefully thank all NAPKON sites who contributed patient data and/or biosamples for this analysis. We also thank the NAPKON Steering Committee: University Hospital Giessen and Marburg, Giessen (Herold S), University of Wuerzburg, Wuerzburg (Heuschmann P), Charité - Universitätsmedizin Berlin, Berlin (Heyder R), University Medicine Greifswald, Greifswald (Hoffmann W), Hannover Unified Biobank, Hannover Medical School, Hannover (Illig T), University Hospital Schleswig-Holstein, Kiel (Schreiber S), University Hospital Cologne and University Hospital Frankfurt, Cologne and Frankfurt (Vehreschild JJ), Jena University Hospital, Jena (von Lilienfeld-Toal M), Charité - Universitaetsmedizin Berlin, Berlin (Witzenrath M).

We would like to thank the Core Unit for Confocal Microscopy and Flow Cytometry-based Cell Sorting of the IZKF Würzburg for supporting the study. We thank Uta Behrends, Technische Universität München (TUM) for kindly providing ME/CFS and healthy control samples for this study. We thank Timo Ludwig for providing demographics from STAAB database.

## Author Contribution

BKP conceived the idea, developed and supervised the project. ZL, CH, AG, AK carried out majority of experiments; IMP carried out autophagosome work; SK carried out multivariate and other statistical data analysis; AH, CN, VC, BA, TB, SC, JJV, OM, CS, KL, KT, JR, FE, LS, PUH, SS, CM, FS, CS, NB, SK, GR contributed with patient recruitment, sample collection, patient data management; MEA and MW contributed with important reagents and experiments; AS and SL carried out mass spectrometry studies and related data analysis, RN supervised mitochondrial experimental work, analyzed the data. BKP drafted the manuscript with help from MEA. All other authors critically revised the manuscript. All authors approved the submitted version.

## Materials availability

Further information and requests for resources and reagents should be directed to Bhupesh K Prusty.

## Data availability

The mass spectrometry proteomics data for the serum immunoglobulin proteome study have been deposited to the ProteomeXchange Consortium via the PRIDE partner repository with the dataset identifier PXD041945 and the same for HEK293 dUTPase co-IP proteome study with the dataset identifier PXD041942. All the experimental data are deposited to Mendley (doi: 10.17632/4xkft5g9r5.1).

---

**Supplementary Table 1:** Patient demographics.

**Supplementary Table 2:** List of antigens used in the microarray study.

**Supplementary Table 3:** List of plasmids used in the study.

**Supplementary Table 4:** List of antibodies used in the study.

**Supplementary Table 5:** Inter- and Intra-assay variations in IgM and IgG ELISA.

## Online-only Methods

### Patient recruitment and serum collection

#### Long COVID patient cohort

COVIDOM is a prospective, population-based cohort study to investigate the long-term health sequelae of SARS-CoV-2 infection. Details have been reported elsewhere^1, 2^. COVIDOM was approved by the local ethics committees of the university hospitals of Kiel (No. D 537/20), and Würzburg (No. 236/20_z) and was registered at clinicaltrials.gov (NCT04679584) and at the German Registry for Clinical Studies (DRKS, DRKS00023742). COVIDOM participants of the Würzburg sub-cohort were recruited in the catchment area of Würzburg and identified through local public health authorities so as to address an unbiased subpopulation regarding age, sex, hospitalization, and media literacy. Main inclusion criteria were (i) a polymerase chain reaction (PCR) confirmed SARS-CoV-2 infection and (ii) a period of at least 6 months between the infection and the visit to the COVIDOM study site. During the on-site examination participants underwent detailed cardio-pulmonary phenotyping and blood was drawn. Serum was stored at Interdisciplinary Bank of Biomaterials and Data of the University Hospital of Würzburg (ibdw). Long COVID severity was determined on the basis of a previously described scoring system^1^.

#### ME/CFS patient cohort

ME/CFS patients, and gender- and age matched healthy controls were diagnosed at the outpatient clinic at the Charité Universitätsmedizin, Berlin and Technische Universität München (TUM) between 2020 and 2023. Diagnosis of ME/CFS in all patients was based on the 2003 Canadian Consensus Criteria and exclusion of other medical or neurological diseases that may cause fatigue by a comprehensive clinical and laboratory evaluation. The study was approved by the Ethics Committee of Charité Universitätsmedizin Berlin (EA2/067/20; EA2_066_22; EA4_174_22) and TUM (112/14 (healthy controls) and 529/18 (pediatric ME/CFS), 485/18 adult ME/CFS) in accordance with the 1964 Declaration of Helsinki and its later amendments. Some of the patient samples were also collected at University of California San Diego with signed informed consent under UCSD IRB Project #140072, “The UCSD Metabolomics Study”. All patients gave informed consent. Whole blood samples from each subject were allowed to clot at room temperature and then centrifuged at 2000 x g for 10 min. The serum was stored in aliquots at −80°C.

#### Healthy controls

For long COVID studies, additional healthy controls were used from the population-based Characteristics and Course of Heart Failure Stages A-B and Determinants of Progression (STAAB) Cohort that recruited individuals without self-reported HF from the general population of Würzburg, Germany, aged 30-79 years and stratified for age and sex. The detailed study design and methodology have been published^3^. All study related procedures were subjected to a rigid and regular quality control process^4^. The STAAB cohort study protocol and procedures comply with the Declaration of Helsinki and received positive votes from the Ethics Committee of the Medical Faculty as well as from the data protection officer of the University of Würzburg (vote #98/13). All participants provided written informed consent prior to any study examination. During the baseline visit, a fasting blood sample was taken and serum was stored in the Interdisciplinary Bank of Biomaterials and Data of the University Hospital of Würzburg (ibdw). For the current project, we identified a random cohort of samples of STAAB participants, between the age 40 and 79 years. Overall characteristics of the patient cohort is summarized in Supplementary Table 1.

For all the above-mentioned patient and healthy control study groups, there were no statistically significant differences in age between male and female subjects except for healthy controls (p = 0.018).

### Cell culture

U2-OS (HTB-96), HEK293 (CRL-1573) cells were purchased from ATCC and HUVEC-TERT2 cells (CHT-006-0008) were purchased from Evercyte, Austria. U2-OS and HEK293 cells were grown in McCoy’s 5A and DMEM media respectively supplemented with 10% (v/v) FBS and 200 units/ml penicillin-streptomycin. HUVEC-TERT2 cells were grown in EBM basal medium (Lonza, Cat#CC-3121) supplemented with Components of EGM SingleQuot Kit (Lonza, Cat# CC-4133: BBE, hEGF, hydrocortisone, ascorbic acid), 10% FBS and 20 µg/ml G418. All cell lines were cultured at 37 °C with 5% CO_2_. Cells carrying stable GFP expression within mitochondria were developed as mentioned before^5^. Cells stably expressing the mitochondrial targeted GFP were created by cloning the mitochondrial targeted GFP into pLVTHM vector backbone (Supplementary Table 3) and transducing target cells with the lentivirus as mentioned before^5^. All the cell lines were frequently tested for Mycoplasma contamination and were authenticated by sequencing, wherever necessary.

### dUTPase constructs, recombinant protein purification

Constructs used in this study are detailed in Supplementary Table 3. Briefly, HSV-1 dUTPase (UL50), HHV-6A dUTPase (U45) and EBV dUTPase (BLLF3) were cloned from bacterial artificial chromosome (BAC) carrying the full-length viral genome. pFN22A vector backbone was used to clone the viral dUTPases carrying N-terminal Halo-tag. All the constructs were sequenced to confirm absence of mutations within the dUTPase open reading frames. A mock dUTPase was created to be used as experimental control by digesting the pFN22A-halo-HHV-6 dUTPase construct with XbaI restriction enzyme which removed majority of viral dUTPase except for 186 bp N-terminal sequences. Digested construct was religated, sequence verified and used as mock control.

For dUTPase recombinant protein preparation, bacterial Strain BL21 (DE3)plyS: F-, ompT, hsdSB (rB-mB-) gal, dcm with chloramphenicol resistance was used. pTrcHIS vector (Thermo Fischer Scientific) was selected using Ampicillin resistance. Depending on the particular dUTPases constructs, bacterial pellets were obtained from 6 to 54 liters of Luria-Broth (LB) culture. An overnight culture was prepared by inoculation of 100-200 µl of the bacterial strain into 200 ml of LB medium containing ampicillin (100 μg/ml) and chloramphenicol (5 μg/ml). with shaking at 37 °C. Next day, a fresh inoculation was prepared by mixing 1 liter of LB medium containing ampicillin and chloramphenicol with 20-25 ml of the overnight culture and was incubated at 37 °C for 2 h with shaking. IPTG was added to the inoculated medium at a concentration of 24 μg/ml with a further incubation of 2 h with shaking at 37 °C. Bacterial cells were centrifuge at 5000 rpm for 5 min and pellets were frozen down at −20 °C till protein extraction. On the day of protein preparation, thawed cell pellets were mixed in column equilibration buffer (50 mM sodium phosphate, 300 mM NaCl and 10 mM imidazole, pH 7.4), sonicated for 4 min (1 min burst; 1 min rest) on ice, centrifuged at 10,000 rpm for 30 min and the the supernatant was stored at 4 °C till the next day. HisPurTM Cobalt 3 ml spin columns were used under native conditions for protein purification. dUTPase proteins were eluted with elution buffer (50 mM sodium phosphate, 300 mM NaCl and 150 mM imidazole, pH7.4).

### Average mitochondrial surface area and mitochondrial number analysis

Software and modified algorithm for mitochondrial size and number measurement were previously described by us in detail^5, 6^. All image-processing and analysis steps were performed using Fiji^7^.

### Immunofluorescence Microscopy

A detailed protocol for standard immunofluorescence microscopy is previously described^8^. All the antibodies used in the immunofluorescence studies are described in Supplementary Table 4. Halo-tagged dUTPase expression and cellular localization was visualized using Halo-tag TMRDirect ligand (Promega, Cat. G2991).

### Immunoblotting

Immunoblotting was carried out as described before^8, 9^ using antibodies mentioned in Supplementary Table 4. Equal protein loading was confirmed by using antibodies against β-actin or GAPDH. All the primary antibodies were used at a dilution of 1:1000 and the HRP-conjugated secondary antibodies were used at a dilution of 1:10,000.

a. ***Western blot for detecting fibronectin in serum:*** Serum samples were diluted 1:1 by 2X Laemmli buffer (100 mM Tris pH 6.8, 4% SDS, 0.2% bromophenol blue, 20% glycerol and 200 mM β-mercaptoethanol) and then further diluted 1:100 by 1X Laemmli buffer (50 mM Tris pH 6.8, 2% SDS, 0.1% bromophenol blue, 10% glycerol and 100 mM β-mercaptoethanol). 10 μl diluted serum samples were loaded onto 4-10% gradient SDS-PAGE gels. 1 μg of human plasma fibronectin (Sigma-Aldrich, F2006) and 0.1 μg fibronectin fragment 2 (Sino Biological, 10314-H08H) were loaded as control onto the SDS-PAGE gels.

b. ***Western blot for detecting natural IgM and IgG:*** 1 μg of human plasma fibronectin (Sigma-Aldrich, F2006) and 0.1 μg fibronectin fragment 2 (FN1.2) (Sino Biological, 10314-H08H) were loaded onto SDS-PAGE gels. Each blot was incubated with 50 ul of human serum in 5 ml of 3% BSA for overnight at 4 °C. HRP-conjugated anti-human IgG or IgM was used (1:10000 dilution) for detecting the fibronectin bound IgG or IgM.

c. ***Western blot for detecting IgG against herpesvirus dUTPase:*** 5 μg of recombinant EBV, HSV-1 and HHV-6 dUTPase proteins were loaded onto 10% SDS-PAGE gels. After gel running and transfer onto a nitrocellulose membrane, each membrane was incubated with 50 ul of human serum in 5 ml of 3% BSA for overnight at 4 °C. HRP-conjugated anti-human IgG was used (1:10000 dilution) for detecting the IgG against individual herpesvirus dUTPases. Intensities of appropriate bands were quantified using densitometric analysis. An arbitrary grading system (0, 1, 2 and 3) was used to evaluate amounts of IgG response against respective herpesvirus dUTPases. 0, absent (no IgG response detected); 1, low; 2, moderate; 3, high. Specificity and cross-reactivity of individual herpesvirus IgG against each other was checked.

### Analysis of mitochondrial membrane potential by TMRE (Tetramethylrhodamine-ethylester-perchlorate)

For the analysis of mitochondrial membrane potential HEK293 cells were seeded in 6 well plates. Cells were transfected with plasmids encoding Halo-tagged herpesvirus dUTPase (Supplementary Table 3). After 48 hours cells were trypsinized, washed and counted. For TMRE staining (Sigma-Aldrich, 87917) 2 X 10^6^ cells were washed with PBS and resuspended in a 50 nM TMRE solution in PBS and 137 mM KCl and stained for 10 min at room temperature. Cells were measured by flow cytometer (Attune Flow Cytometers) recording first base level of TMRE fluorescence. Then, half of the cells were treated with 1.5 µM of Oligomycin (Sigma Aldrich, #04876 in DMSO). Fluorescence was measured at different timepoints of incubation. At the end, 5 µM FCCP (Carbonyl cyanide-p-trifluoro­methoxy­phenyl­hydra­zone, Sigma Aldrich, #C2920) was added to induce mitochondrial depolarization and the changes in fluorescence levels because of loss of membrane potential was measured.

### Immunoglobulin purification

150 µl Protein G Sepharose 4 Fast Flow (#17061801,cytiva) was loaded onto Poly-Prep Chromatography Columns (#731-1550, BIO-RAD). After washing the beads with PBS, 500 µl serum sample was loaded and passed through the column 3 times, which was followed by 3 time washing by PBS. Protein G bounded serum IgG was eluted by Glycine pH 2.7 and then neutralised with 1M Tris-HCl pH 8.0. IgG elute was dialyzed against PBS in Slide-A-Lyzer MINI Dialysis Devices (#88404, Thermo Fisher Scientific). IgG concentration was measured by Easy-Titer Human IgG (H+L) Assay Kit (#23310, Thermo Fisher Scientific).

### Co-immunoprecipitation of His-tagged viral dUTPases

Halo-tagged herpesvirus dUTPase immunoprecipitation (IP) was carried out using standard IP protocol^5^ using ysis buffer (50 mM Hepes, pH 7,5; 150 mM NaCl; 0.5% NP-40; 1 mM NaF; 10% Glycerol, 2.5 mM MgCl_2_; protease inhibitor cocktail (Roche), 0.5 mM DTT) and wash buffer (50 mM Hepes, pH 7,5; 150 mM NaCl; 1 mM NaF; 10% Glycerol, 2.5 mM MgCl_2_; protease inhibitor cocktail (Roche), 0.5 mM DTT).

### IgG exposure in cell culture

U2-OS or HUVEV-TERT2 cells carrying soluble mitoGFP were seeded on 6-well plates and cultured overnight. Cell culture medium containing 1 µg/ml serum IgG was added to each well, allowing cells to be exposed to serum derived IgG. Cells were collected after 24 h for western blot or immunofluorescence experiments.

### Fibronectin ELISA

Human fibronectin ELISA was performed with Fibronectin Human ELISA Kit (#BMS2028, Thermo Fisher Scientific) by the manufacturer’s protocol.

### IgM-FN1 and IgG-FN1 ELISA

96 well microplates (Sigma-Aldrich, CLS9018) were coated with 100 ng fibronectin antibody (Santa Cruz, sc-18825) in carbonate coating buffer (Thermo Fisher Scientific, CB01100) overnight at 4°C. For IgG and IgM standard wells, 100 μl 25.60 ∼ 0 pg/μl series dilutions of native human IgM protein (Abcam, ab90348) or human IgG isotype control (Thermo Fisher Scientific, 31154) was coated onto the plates in carbonate coating buffer (Thermo Fisher Scientific, CB01100) overnight at 4°C. Plates were then washed 3 times for 5 min by 200 μl 1X wash buffer on a plate shaker, which was prepared by diluting 20X TBST (Thermo Fisher Scientific, 28360) in distilled water. 2% BSA (Bovine Serum Albumin) (Sigma-Aldrich, A2153) was dissolved in 1X washing buffer. 300 μl 2% BSA were used to block the plates for 1h at RT and the plates were washed 1 time afterwards. Serum samples from patients and healthy control were diluted 1:100 by 2% BSA. Plates were then incubated with 100 μl diluted serum samples in each well for 1 h at RT on a shaker. Plates were then washed 6 times for 5 min by 200 μl 1x washing buffer on a shaker. Goat anti-human IgM-HRP (Thermo Fisher Scientific, 31415) and rabbit anti-human IgG-HRP (Sigma-Aldrich, A8792) were used as secondary antibodies for signal detection. Secondary antibodies were diluted 1:10000 in 2% BSA. Plates were incubated with 100 μl of diluted secondary antibody for 1h at RT and then washed 6 times for 5 min by 200 μl 1X washing buffer on a plate shaker. HRP signals were developed by incubating the plates with 100 μl TMB substrate solution (Thermo Fisher Scientific, 34029) for 15-30 minutes at RT until desired color was visible and then stopping the reaction by adding 100 μl stop solution (Thermo Fisher Scientific, SS04). Plates were immediately read by an optical plate reader (Avantor, SpectraMax PLUS384) at 450 nm. The concentration of the samples was calculated by using the standard curves as references. Duplicates were applied for all unknown samples and standard samples.

#### Principles of sandwich ELISA

Fibronectin antibody (Santa Cruz, sc-18825) was raised against the cell binding domain, which is located within the FN1.2 domain. However, natural IgM against fibronectin only showed binding activity with plasma fibronectin instead of recombinant FN1.2, suggesting that natural IgM against fibronectin and fibronectin antibody (Santa Cruz, sc-18825) have different binding domain. Based on this, we developed our sandwich ELISA for IgM and IgG.

#### Inter- and Intra-assay validation for IgM-FN1 and IgG-FN1

For inter- and intra-assay accuracy testing, optical density (O.D.) values at 450 nm for standard concentration curves between the range of 0.8 ng/ml to 25.6 ng/ml human native IgM and human IgG controls were generated (Supplementary Table 5a and 5b respectively). Two independent experiments on three different days (Six independent standard curves in total), for both IgM and IgG, were developed and carried out. Intra-assay precision was assessed over these six independent standard concentration curves. Each standard concentration was performed in duplicates. The results showed overall intra-assay coefficients of variation (CV) of 3.45% (range between 0.00% to 14.93%) for IgM and 2.67% (range between 0.00% to 8.62%) for IgG. Inter-assay reproducibility was determined by the overall CV of all mean O.D. values from six independent experiments. The results showed CV of 3.22% (range between 0.02% to 11.47%) for IgM and 3.23% (range between 0.03% to 11.48%) for IgG.

### Mass spectrometry

A fraction of serum IgG bounded protein G beads from IgG purification was eluted by NuPAGE LDS Sample Buffer (4X) (#NP0007, Thermo Fisher Scientific). The elutes were sent for mass spectrometry.

### Single-pot, solid-phase-enhanced sample preparation (SP3)

Samples were processed using an adapted SP3 protocol^10^. Briefly, 200 µl reconstitution solution was added to each sample prepared in 50 µl NuPAGE LDS sample buffer (Life Technologies). Reduction was performed using 5 mM DTT with subsequent alkylation with 20 mM iodoacetamide. 10mM additional DTT was used for quenching. Equal volumes of two types of Sera-Mag Speed Beads (Cytiva, #45152101010250 and #65152105050250) were combined, washed with water and 10 µL of the bead mix were added to each sample. 260 µl 100% ethanol was added and samples were incubated for 5 min at 24 °C, 1000 rpm. Beads were captured on a magnetic rack for 2 min, and the supernatant was removed. Beads were washed twice with 200 µl 80% ethanol (Chromasolv, Sigma) and then once with 1000 µl 80% ethanol. Digestion was performed on beads with 0.25 µg Trypsin (Gold, Mass Spectrometry Grade, Promega) and 0.25 µg Lys-C (Wako) in 100 µl 100 mM ammonium bicarbonate at 37 °C overnight. Peptides were desalted using C-18 Stage Tips^11^. Each Stage Tip was prepared with three discs of C-18 Empore SPE Discs (3 M) in a 200 µl pipet tip. Peptides were eluted with 60 % acetonitrile in 0.1 % formic acid, dried in a vacuum concentrator (Eppendorf), and stored at −20 °C. Peptides were dissolved in 2 % acetonitrile / 0.1 % formic acid prior to nanoLC-MS/MS analysis.

### NanoLC-MS/MS analysis

NanoLC-MS/MS analyses were performed on an Orbitrap Fusion (Thermo Scientific) equipped with a PicoView Ion Source (New Objective) and coupled to an EASY-nLC 1000 (Thermo Scientific). Peptides were loaded on a trapping column (2 cm x 150 µm ID, PepSep) and separated on a capillary column (30 cm x 150 µm ID, PepSep) both packed with 1.9 µm C18 ReproSil and separated with a 120-minute linear gradient from 3% to 30% acetonitrile and 0.1% formic acid and a flow rate of 500 nl/min. Both MS and MS/MS scans were acquired in the Orbitrap analyzer with a resolution of 60,000 for MS scans and 30,000 for MS/MS scans. HCD fragmentation with 35 % normalized collision energy was applied. A Top Speed data-dependent MS/MS method with a fixed cycle time of 3 s was used. Dynamic exclusion was applied with a repeat count of 1 and an exclusion duration of 90 s; singly charged precursors were excluded from selection. Minimum signal threshold for precursor selection was set to 50,000. Predictive AGC was used with AGC a target value of 4×10^5^ for MS scans and 5×10^4^ for MS/MS scans. EASY-IC was used for internal calibration.

### MS data analysis

Raw MS data files were analyzed with MaxQuant version 1.6.2.2^12^. Database search was performed with Andromeda, which is integrated in the utilized version of MaxQuant. The search was performed against the UniProt Human Reference Proteome database (June 2022, UP000005640, 79684 entries), the UniProt HHV-6 Reference Proteome database (October 2018, UP000009295, 107 entries) and the UniProt Epstein-Barr virus database (September 2022, UP000272970, 82 entries). For dUTPase co-IP experiment, a small database containing the sequence of the Halo-Tag (UniProt DHAA_RHORH) and the dUTPases of interest (UniProt L0N6A5, P30007, P03195) was used. Additionally, a database containing common contaminants was used. The search was performed with tryptic cleavage specificity with 3 allowed miscleavages. Protein identification was under control of the false-discovery rate (FDR; <1% FDR on protein and peptide spectrum match (PSM) level). In addition to MaxQuant default settings, the search was performed against following variable modifications: Protein N-terminal acetylation, Gln to pyro-Glu formation (N-term. Gln) and oxidation (Met). Carbamidomethyl (Cys) was set as fixed modification. LFQ intensities were used for protein quantitation^13^.

For Halo-tag co-IP data analysis, proteins with less than two razor/unique peptides were removed. Missing LFQ intensities were imputed with values close to the baseline. Data imputation was performed with values from a standard normal distribution with a mean of the 5% quantile of the combined log10-transformed LFQ intensities and a standard deviation of 0.1. For the identification of significantly enriched proteins, boxplot outliers were identified in intensity bins of at least 300 proteins. Log2 transformed protein ratios of sample versus control with values outside a 1.5x (significance 1) or 3x (significance 2) interquartile range (IQR), respectively, were considered as significantly enriched.

### Antigen Microarray experimental set up

Microarray studies for IgM against autoantigens were carried out in collaboration with Creative Biolabs, USA. Frozen serum samples without prior freeze-thaw cycle were used for the assay. Each serum was digested with DNase I for 30 min at room temperature on a shaker. For the control, no serum sample was added. Slides carrying antigens against 120 autoantigens (for details of the antigens, please see Supplementary Table 2) were blocked in 100 ml blocking buffer at room temp for 30 min on a shaker. Afterwards slides were washed twice with PBST each for 5 min. 90 ml PBST was added into each serum sample or control mix. Diluted samples were added into each well of the slide (100 ml /each) and were incubate at room temp for 1 hour on a shaker. Slides were washed with PBST 100 ml/well for 5 min on the shaker. Subsequently slides were washed with blocking buffer 100 ml/each well for 5 min on a shaker and the with PBST 100 ml/each well, 5 min on a shaker. Anti-human IgM secondary antibody were diluted to 1:1000 in PBST and 100 ml secondary antibody was added to each well. Slides were incubated at room temp for 1 hour on the shaker, washed three times with PBST 100 ml/each well, 5 min on the shaker. Slides were then washed twice each, first with 45 ml PBS in 50 ml tube for 5 min on the shaker and then with 45 ml nuclease-free water in 50 ml tube for 5 min on the shaker. GenePix 4000B microarray systems was used to scan the slide. 532 nm channel was used to scan Cy3 fluorescence, and 635 nm channel was used to scan Alexa Fluor-647 fluorescence.

### Microarray data analysis

Each chip had serially diluted anti-IgM and IgM as positive control to monitor the experimental process, and PBS was used as a negative control. For the obtained chip image, LuxScan 3.0 software was used to read the original data. Statistical tests were then carried out on the chip background, the signal intensity of the positive and negative control sites. The results showed that the Ig control signal value was higher, and the uniformity was good in different samples. In addition, the value of PBS anti-Ig control and background signal value were all low, which met the quality control requirements. The chip of the test sample was scanned with LuxScan 10K-B scanner. Autoantigen microarray chip/ Pathogen-associated antigen microarray chip had 256 points in total. After removal of the 8 anti-IgM and 8 Ig control points, 240 data points were finally obtained. Each protein on the chip was present as two technical repetitions, representing 120 autoantigens. The chip is read by LuxScan 3.0 software to obtain the original data, including foreground signal (F Median), background signal (B Median) and so on. Foreground Median, Background Median columns were extracted from the .LSR file. Fluorescence intensity value of each site was calculated by the formula: Net Fluorescence Intensity (NFI) Value = (Foreground Median - Background Median); SNR = (Foreground Median - Background Median) / SD(Background). SNR was used to filter unreasonable data. The net fluorescence value was set as SNR <0.05 and SNR to 0.001. The net fluorescence value was calculated after subtracting the blank control PBS NFI_No PBS_. The NFI and SNR of the following unreasonable situations was set to 0.001. NFI<20 and SNR>5; SNR<0.05 and NFI>20; NFI<0.05. RLM Normalization was used to normalize the NFI and calculate the effect values of different blocks and slides. For the microarray data clustering and multivariate analysis, first log transformation was carried out and then ‘pheatmap’ was used using R package with Ward.D2 clustering to make the cluster analysis.

### Machine learning and Multivariate analysis of autoantigen IgM microarray data

Multivariate analysis of patient clusters based on distance metrics derived from IgM antibody levels against for a panel of autoantigens. Columns with zero variation (constant values) were removed. Log-transformed scaled data showing relative differences between patients was used because the data in its raw form had different orders of magnitude, making analysis and comparison difficult. The analysis was performed using the R package “pheatmap” using Ward.D2 clustering method and Euclidean distances.

A RandomForest classifier using the R package randomForest was fit predicting ME/CFS as an outcome was performed on two types of data - raw data and log scaled data. The purpose of this fit was a rapid screen for variables for further analysis. The Variable Importance Plot for the fits were plotted. The top 10 candidates from the Log Variable Importance Plots were selected for Principal Component Analysis (PCA), a statistical methodology for dimension reduction. PCA was was performed using the R package stats (prcomp) and FactoMineR.

### Statistical testing of herpesvirus dUTPase antibody

As a part of the Likert Analysis, the following steps were performed: Counts and percentage of totals were generated in R using dplyr package. The above results were grouped into relevant sets (EBV, HSV-1, HHV-6, Healthy, ME/CFS, Covid-19 PCR positive but no long COVID (No LC), Mild LC, Severe LC). Likert charts were generated from the above groupings.

Non-parametric analysis was performed because the groups were of different sizes. The R package “rcompanion” was used because it has the higher order non-parametric ScheirerRayHare Test. Kruskall Wallis and its associated higher order test ScheirerRayHare Test were performed because two factors (Antibody State, i.e., 0, 1, 2, 3 and virus state, i.e., EBV, HSV-1, HHV-6) are changing simultaneously and it would be useful to know which factor was statistically significant. For several tests, it appeared that Antibody State was significant.

### Other statistics

Kernel density plot showing the bivariate serum FN1 distributions among healthy controls and ME/CFS patients was performed using the Python package package Seaborn *(Seaborn.kdeplot*) . All other statistical calculations were performed using GraphPad Prism 9.0. Error bars displayed on graphs represent the means ± SD of three or more independent replicates of an experiment. Statistical significance was calculated separately for each experiment and are described within individual figure legends. For image analysis, three or more biological replicates per sample-condition were used to generate the represented data. The results were considered significant at P ≤ 0.05.

### Reporting summary

Further information on research design is available in the Nature Research Reporting Summary linked to this paper.

### Data and Materials availability

Further information and requests for resources and reagents should be directed to Bhupesh K Prusty. The mass spectrometry proteomics data for the serum immunoglobulin proteome study have been deposited to the ProteomeXchange Consortium via the PRIDE partner repository with the dataset identifier PXD041945 and the same for HEK293 dUTPase co-IP proteome study with the dataset identifier PXD041942. All the experimental data are deposited to Mendley (doi: 10.17632/4xkft5g9r5.1).

## Extended Figure legends

**Extended Fig. 1:**
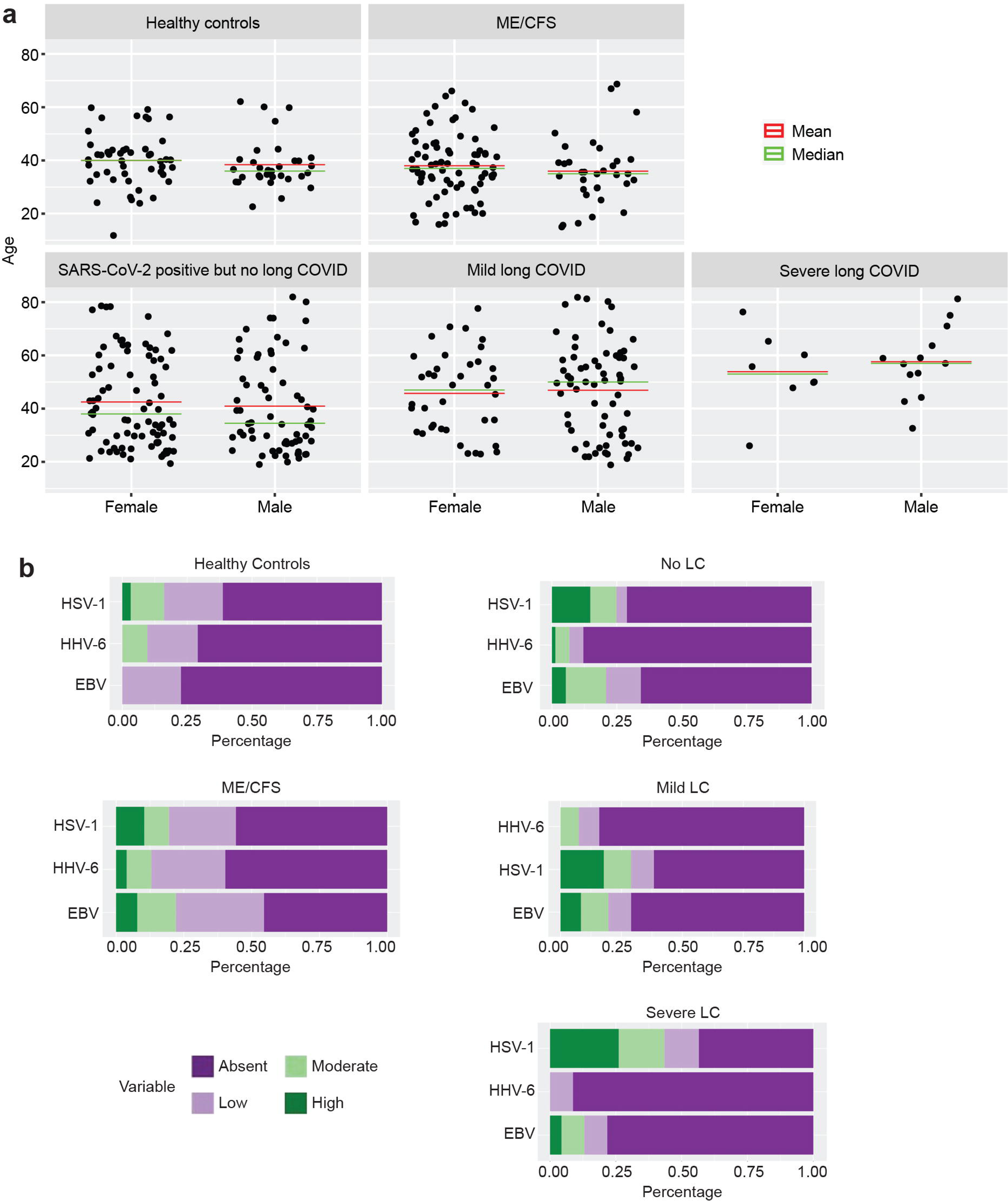
Herpesvirus reactivation in healthy population, ME/CFS, long COVID patients. **a.** Demography of distribution of patients according to the age and gender. **b.** Likert chart showing percentage of positivity for antibodies against EBV, HSV-1 and HHV-6 dUTPase within healthy controls (HC), ME/CFS, Covid-19 PCR positive but without long COVID (No LC), mild LC and severe LC patients. Kruskall Wallis (and its higher order equivalent, ScheirerRayHare Test) rank sum test for antibody state. HC, *P = 0.019; ME/CFS, *P = 0.021. Different amounts IgG levels in patient serum was arbitrarily divided into 4 groups (0, absent; 1, low; 2, moderate; 3, high).

**Extended Fig. 2:**
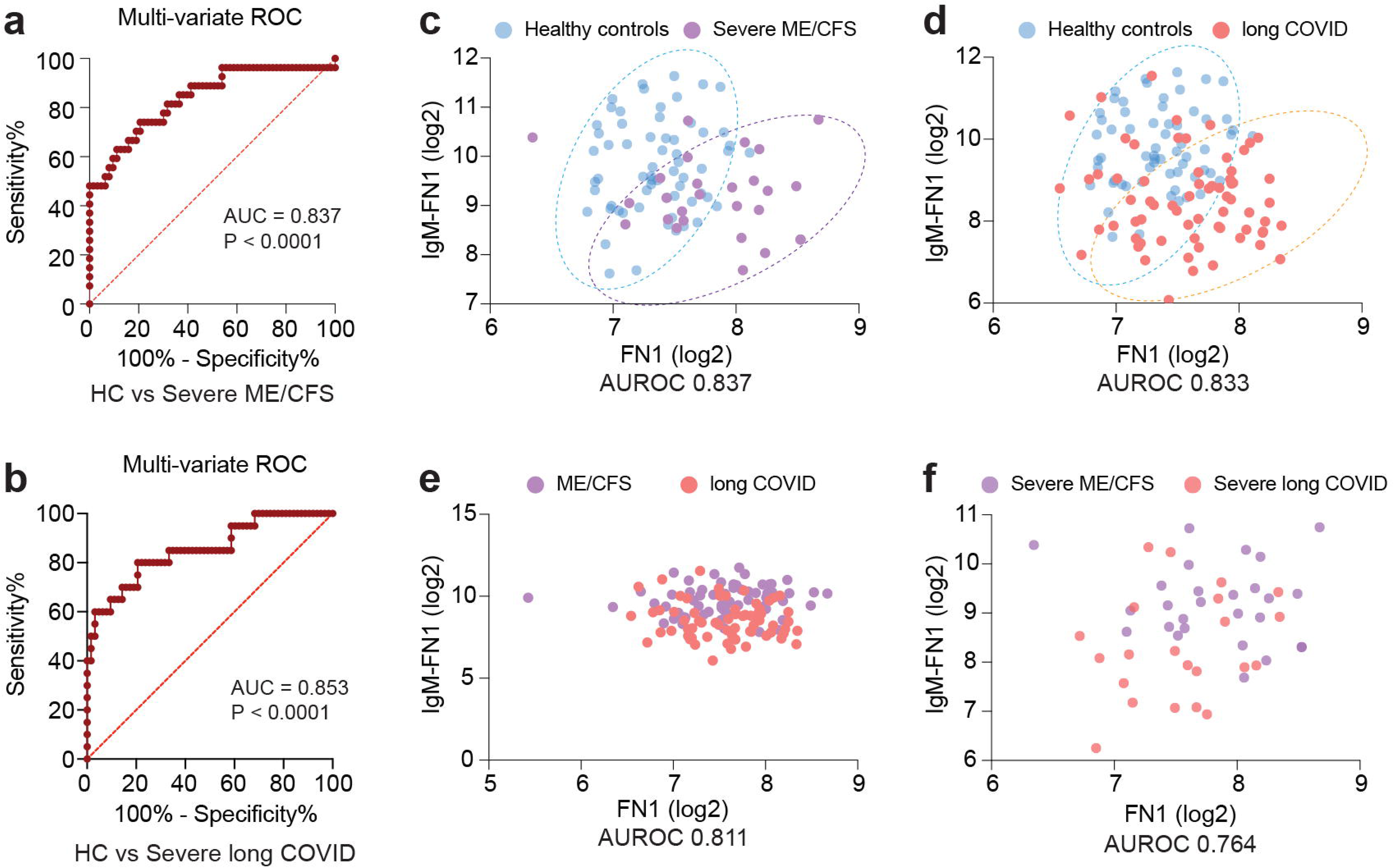
Role of viral dUTPase proteins in mitochondrial dysfunction. **a**. Confocal images showing hyperpolarization of mitochondria in U2-OS cells under transient expression of HSV-1, HHV-6 and EBV dUTPases. Mock vector was used as a control. **b**. Average mitochondrial surface area was compared between mock vector and EBV dUTPase transfected U2-OS cells. Data from three independent biological replicates. n=3. Unpaired two tailed non-parametric t-test. *P = 0.0278. **c.** Immunoblot analysis shows no major changes in dynamin related protein 1 (Drp1), p53, mitofusin1 (Mfn2), mitoguardin1 (Miga1) protein levels in presence of herpesvirus dUTPases. GAPDH staining was used as a loading control. **d.** TMRE dye was used to study mitochondrial membrane potential and OXPHOS in HEK293 cells. Cells were transiently transfected with HHV-6, HSV-1 dUTPases or a mock vector for 48 h. Non-transfected cells are also used as a control. Trypsinized cells were stained with TMRE dye and were used for flow cytometry. Oligomycin was used to inhibit ATP synthase. Data from 3 independent experiments. n=3. **e.** Data from above experiment was used to compare hyperpolarization status of mitochondria between mock transfected and EBV dUTPase transfected cells. Data from 3 independent experiments. n=3. **f.** Confocal images showing fragmentation of mitochondria in U2-OS cells in presence of recombinant EBV dUTPases. Same amounts of heat inactivated recombinant protein was used as a control. Average mitochondrial surface area was compared between control and recombinant EBV dUTPase exposed U2-OS cells. Data from four independent biological replicates. n=4. Unpaired two tailed non-parametric t-test. **P = 0.0032. **g.** Immunoblot analysis shows upregulation of dynamin related protein 1 (Drp1) levels in U2-OS cells exposed to recombinant EBV dUTPase for 24 h in a dose dependent manner. GAPDH staining was used as a loading control. Drp1 levels are quantified from three independent biological replicates and are shown in the form of a scatter plot. n=3. Unpaired two tailed non-parametric t-test. 0 vs 2 μg, *P = 0.0229; 0 vs 5 μg, *P = 0.0360; 0 vs 10 μg, *P = 0.0201. **h.** TMRE dye was used to study mitochondrial membrane potential and OXPHOS in HEK293 cells exposed to recombinant EBV dUTPase. Trypsinized cells were stained with TMRE dye and were used for flow cytometry. Oligomycin was used to inhibit ATP synthase. Data from 3 independent experiments. n=3. **i.** Coomassie dye stained polyacrylamide gel with input and immunoprecipitated samples shows efficient pull down of Halo-tagged herpesvirus dUTPases. Potential protein bands co-immunoprecipitated along with EBV and HSV-1 dUTPase protein is indicated. **j.** Normalized log2 ratio of LFQ (label-free quantitation) intensities of proteins. Fold change of proteins in EBV vs Mock was plotted against the same in HHV-6 vs Mock to highlight proteins that were specifically enriched in EBV dUTPase expressing co-IP. **k.** Normalized log2 ratio of LFQ (label-free quantitation) intensities of proteins. Fold change of proteins in HSV-1 vs Mock was plotted against the same in HHV-6 vs Mock to highlight proteins that were specifically enriched in HSV-1 dUTPase expressing co-IP. Circles indicate identified cellular proteins; circle size correlates with the number of peptides used for quantification. Significantly enriched proteins that are potential interaction partners of EBV (g) and HSV-1 (h) dUTPases are displayed in red. **l.** Immunoblot analysis to validate potential herpesvirus dUTPase interacting partners identified from co-IP.

**Extended Fig. 3:**
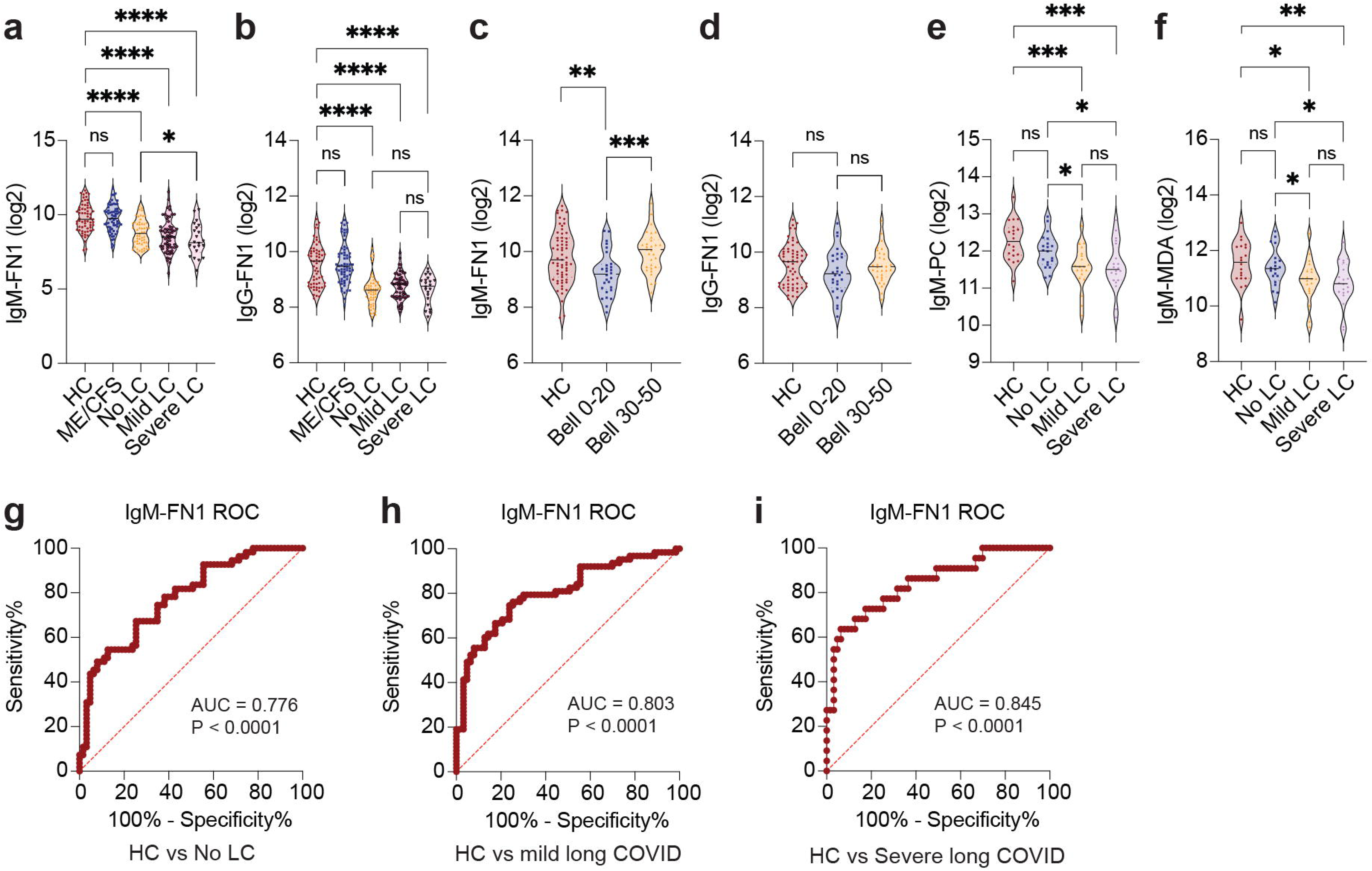
Immune modulations in ME/CFS. **a.** Immunoblot analysis shows validation of enrichment IgG against EBV dUTPase within purified IgG fractions. Recombinant dUTPase proteins are used as a bait. Left panel shows specific detection of EBV dUTPase using an antibody raised against it. Middle panel shows lack of specific signal against EBV dUTPase using a patient serum negative for EBV. Right panel confirms the absence of the specific signal against EBV dUTPase within the purified IgG. GAPDH staining was used as a loading control. **b.** Immunoblot analysis shows positive validation of enrichment IgG against EBV dUTPase within purified IgG fractions. Recombinant dUTPase proteins are used as a bait. Left panel shows specific detection of EBV dUTPase using an antibody raised against it. Middle panel shows specific signal against EBV dUTPases using a EBV positive patient serum. Right panel confirms the presence of the specific signal against EBV dUTPase within the purified IgG. Desired band is indicated with an arrow. GAPDH staining was used as a loading control. **c.** A heat map of proteins identified by mass spectrometry within the purified immunoglobulin complexes. Each row represents a specific protein, and each column represents a specific patient sample. The lower panel of the heat map from upper panel is enlarged in the lower panel along with the protein names. **d.** Log10 values of normalized LFQ intensities of three altered proteins (serotrasferrin, alpha2 microglobulin and fibronectin) comparing healthy controls (n=12) with ME/CFS patients (n=15).

**Extended Fig. 4:**
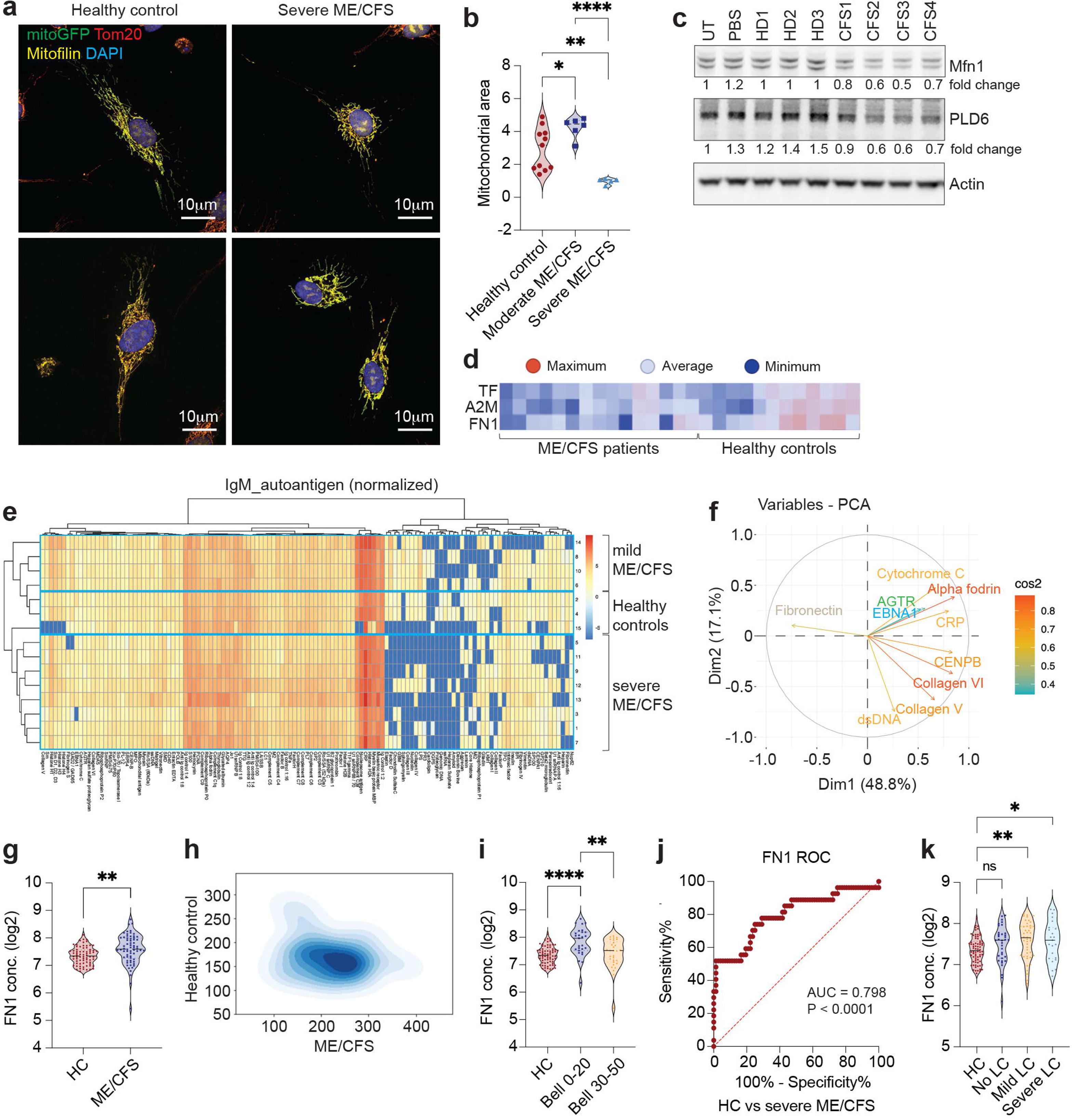
Autoimmune signatures in ME/CFS. **a**. The Variables Factor map (Biplot) for the Principal Components (for patients and healthy Controls combined data) shows the projection of the top 10 Autoantigen variables projected onto the plane spanned by the first two Principal Components. **b**. Biplot of PCA analysis showing variation condensed to key dimensions (dim reduction) for the data set shown in Fig. 2e-f.

**Extended Fig. 5:**
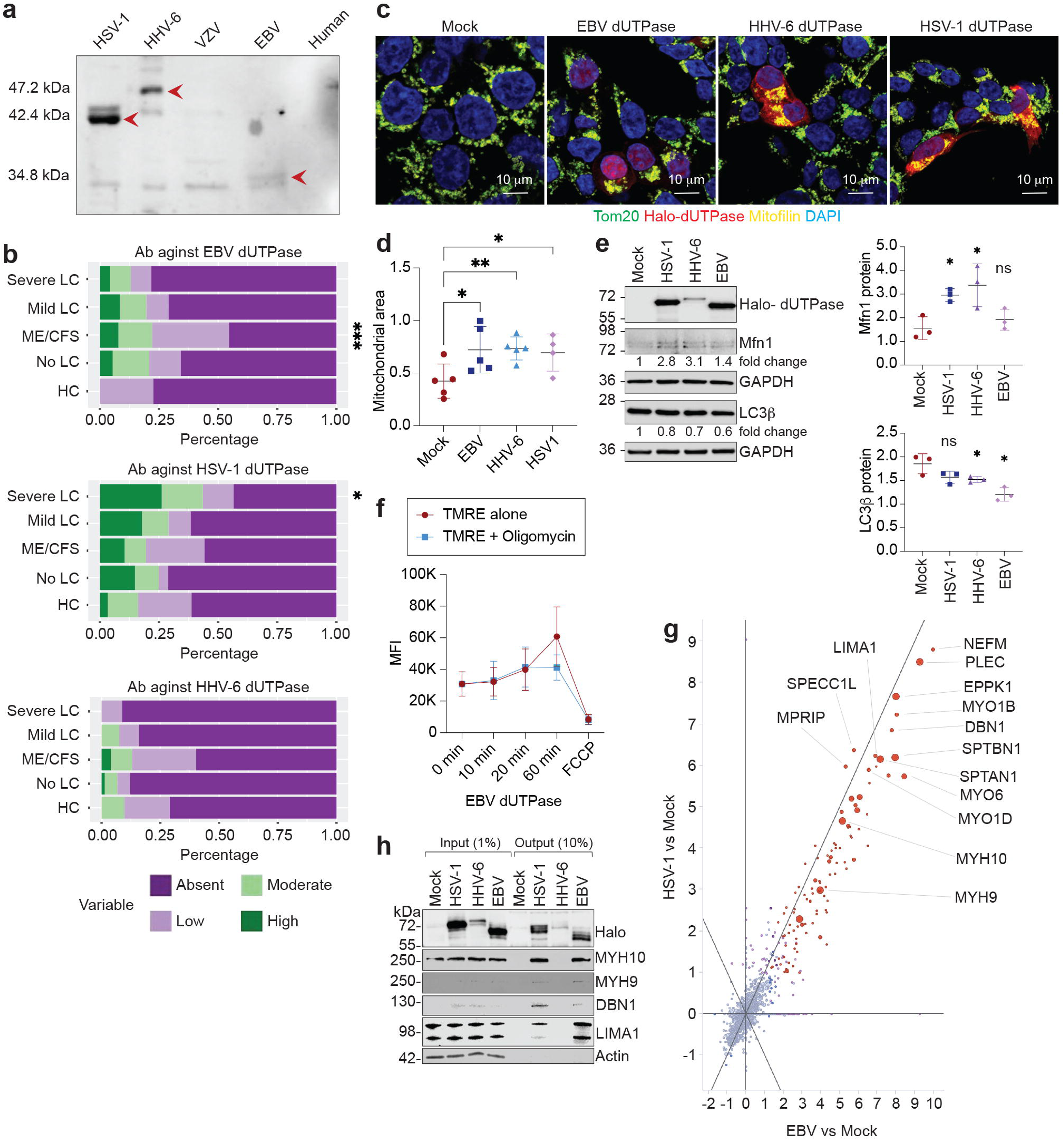
Circulating fibronectin in ME/CFS. **a.** Immunoblot analysis shows overall increase in both plasma fibronectin (plFN1) and cellular fibronectin (clFN1) in ME/CFS patients. Equal amounts of serum proteins from 5 ME/CFS patients with higher serum fibronectin levels were run in parallel with the same from 5 healthy controls. Purified plFN1, recombinant fibronectin isoform FN1.2 and HUVEC cell lysate expressing only cellular fibronectin were used as positive controls. Two different antibodies raised against Extra domain A (EDA) domain and CBD domain of fibronectin was used to differentiate plFN1 from clFN1 as plFN1 lacks EDA domain. **b.** Distribution of circulating FN1 concentrations in different groups of patients separated by gender. Two-tailed parametric t-test. Healthy control (HC) male vs female, **P = 0.0014; No LC male vs female, *P = 0.0204; severe LC male vs female, *P = 0.0347. **c.** Comparison of gender-specific circulating FN1 concentrations (log2 values) among different patient groups. Two-tailed parametric t-test. Healthy control (HC) male vs ME/CFS male, ***P = 0.0001; HC male vs mild LC male, *P = 0.012; HC female vs no LC female, *P = 0.0193; HC female vs mild LC female, ***P = 0.0006; HC female vs severe LC female, **P = 0.0016. **d**. IgM antibody levels against fibronectin (FN1) in ME/CFS patients and healthy controls in the form of a violin plot. IgM levels were determined from microarray studies where each antigen was tested twice separately. Normalized values against signal to noise ratio and net fluorescence intensity is used. **e.** Immunoblot analysis comparing the various species of circulating fibronectin (FN1) proteins in healthy controls and ME/CFS patient sera. Recombinant FN1.2 (recFN1.2) protein lacking extra domain-A (EDA domain) and HEK293 cell lysate serves as control. Three different antibodies detecting specific protein domains (EDA domain, CBD (cell binding domain) and cellular FN (clFN)) of FN1 was used against the same samples to compare the different species of proteins in the sera.

**Extended Fig. 6:**
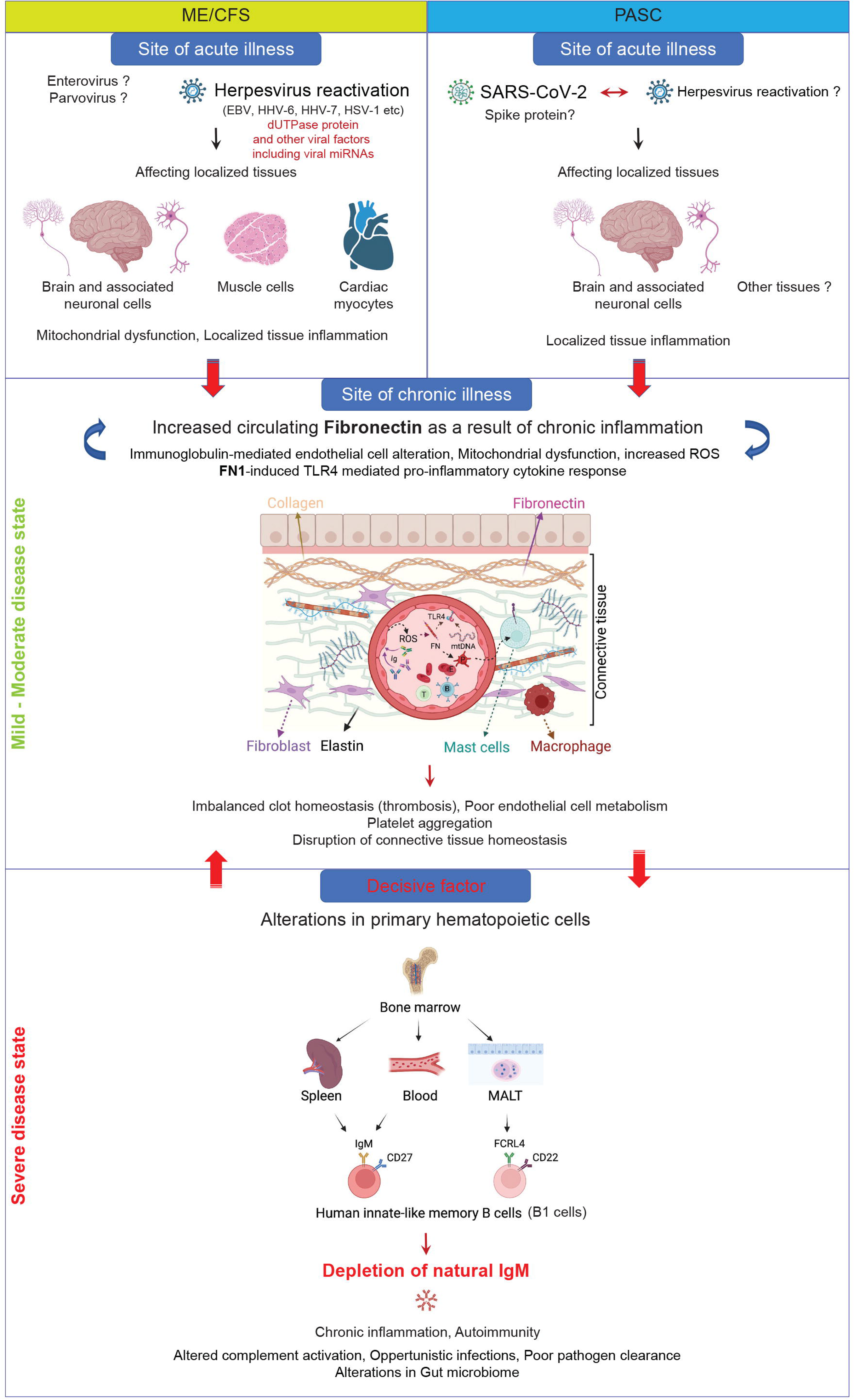
IgM-PC and IgM-MDA distributions among different groups of patients. **a.** Distribution of serum IgM-PC concentrations in different groups of patients separated by gender. **b.** Distribution of serum IgM-MDA concentrations in different groups of patients separated by gender.

**Extended Fig. 7:**
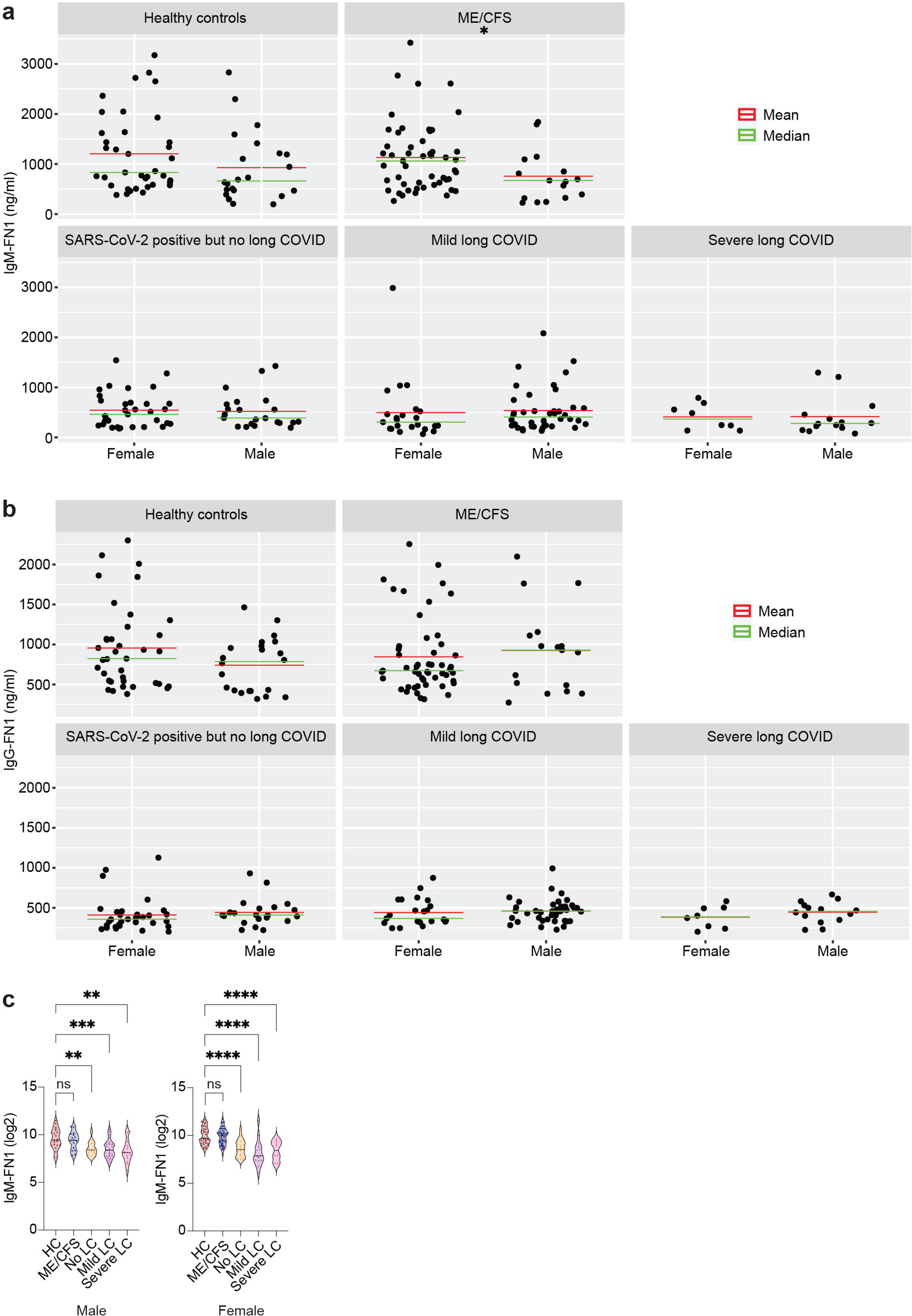
IgM-FN1 and IgG-FN1 distributions among different groups of patients. a. Distribution of serum IgM-FN1 concentrations in different groups of patients separated by gender. Two-tailed parametric t-test. ME/CFS male vs female, *P = 0.0398. **b.** Distribution of serum IgG-FN1 concentrations in different groups of patients separated by gender. **c.** Comparison of gender-specific IgM-FN1 concentrations (log2 values) among different patient groups. Two-tailed parametric t-test. Healthy control (HC) male vs No LC male, **P = 0.003; HC male vs mild LC male, **P = 0.0018; HC male vs severe LC male, ***P = 0.0007; HC female vs no LC female, HC female vs mild LC female, HC female vs severe LC female, ****P < 0.0001.

**Extended Fig. 8:**
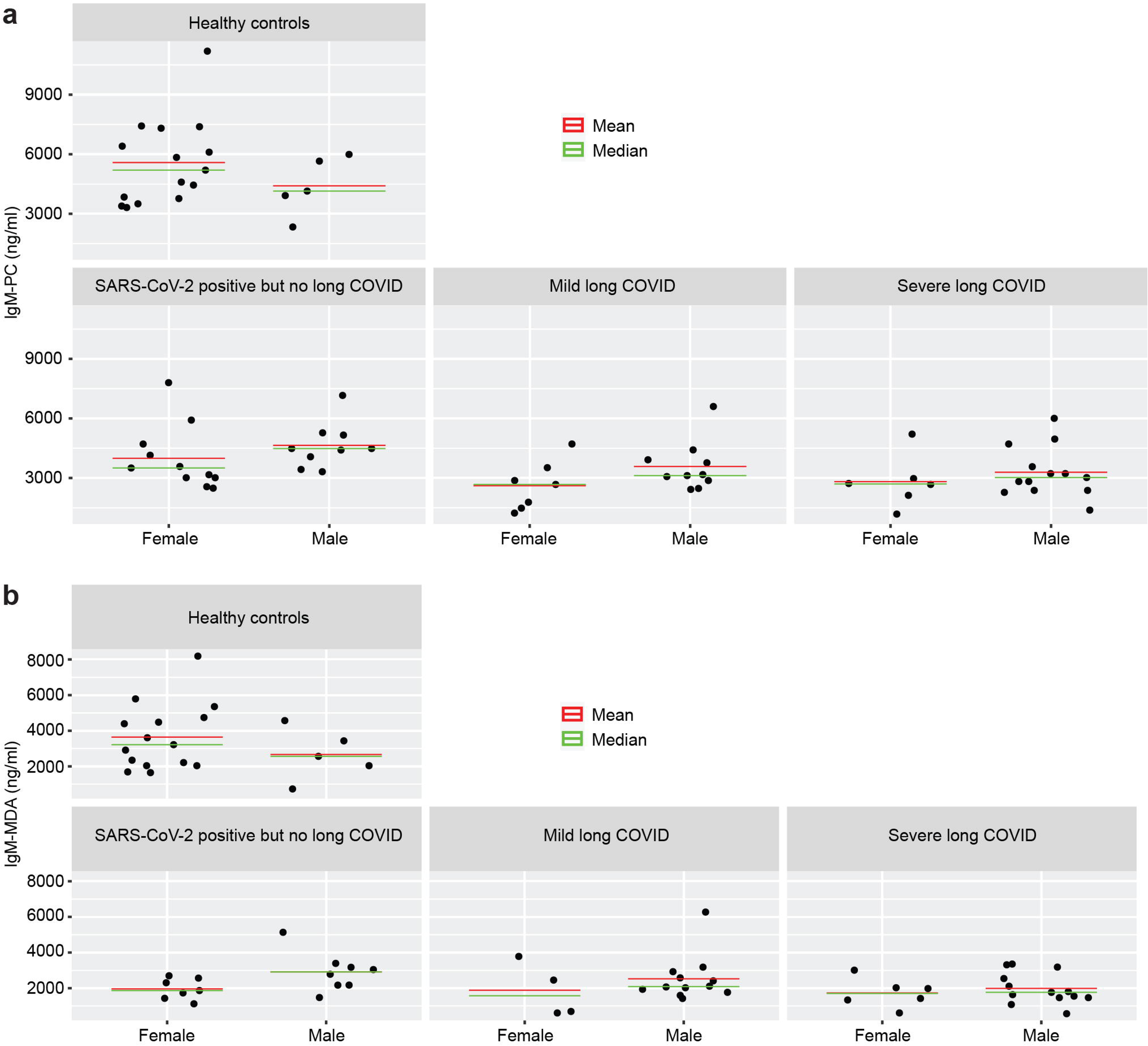
Graphical abstract summarizing potential overlap between ME/CFS and long COVID pathogenesis. Both ME/CFS and long COVID possibly originates as a post viral illness. SARS-CoV-2 infection is the major infection behind long COVID. However, heightened reactivation of herpesviruses like HSV-1 and EBV can potentially play a role in development of ME/CFS. Similar increase in herpesvirus reactivations including those of HSV-1, HHV-6 and EBV are also observed in ME/CFS. Virus-induced direct changes in cellular physiology are expected to be the major driver for the disease development. Subsequently, chronic tissue inflammation could lead to changes in secondary tissue homeostasis where increase in circulating fibronectin levels can play a key role in TLR2/TLR4-mediated innate immune response, cytokine production, Platelet activation, mast cell activation and alterations in clot homeostasis. Major cellular alterations within primary and secondary hematopoietic tissues might lead to substantial decrease in natural IgM production, which subsequently drive the autoimmune feature of both ME/CFS and long COVID. Changed autoimmune signature in the form of autoantibodies could then cause mitochondrial dysfunction, endothelial cell damage initiating a vicious cycle of events that can lead to severe forms of both ME/CFS and long COVID.

