## Supplemental Table 1-5 for "Increased circulating fibronectin, depletion of natural IgM and heightened EBV, HSV-1 reactivation in ME/CFS and long COVID"

| Supplemental Table 1: Patient demographics | | | | | | |
|---|---|---|---|---|---|---|
| | | **Healthy Controls** | **ME/CFS** | **No LC** | **Mild LC** | **Severe LC** |
| n | | 83 | 106 | 149 | 107 | 23 |
| Gender | Female | 51 (61.4%) | 73 (68.9%) | 82 (55%) | 68 (63.5%) | 8 (34.8%) |
| | Male | 32 (38.6%) | 33 (31.1%) | 67 (45%) | 39 (36.5%) | 15 (65.2%) |
| Age | All | 39.3±9.4 | 37.4±12.2 | 41.8±17.1 | 46.6±16.6 | 56±13 |
| | Female | 39.7±9.6 | 38±11.9 | 42.5±16.8 | 46.6±15.1 | 53.9±57.2 |
| | Male | 38.4±9 | 36±12.6 | 40.9±17.3 | 47.1±17.4 | 13.7±12.4 |

| Supplemental Table 2 |
|---|
| **Autoantigens** |
| Aggrecan |
| AGTR |
| Alpha Fodrin |
| alpha-actinine |
| Amyloid |
| AQP4 |
| B2 glycoprotein 1 |
| B2-microglobulin |
| BPI |
| Cardolipin |
| CD40 |
| CENP-A |
| CENP-B |
| Chondroitin Sulfate C |
| Collagen I |
| Collagen II |
| Collagen III |
| Collagen IV |
| Collagen V |
| Collagen VI |
| complement C1q |
| complement C3 |
| complement C3a |
| complement C4 |
| complement C5 |
| complement C6 |
| complement C7 |
| complement C8 |
| complement C9 |
| Core Histone |
| CRP |
| Cytochrome C |
| Decorin-bovine |
| DGPS |
| DNA Polymerase beta (POLB) |

| |
|---|
| dsDNA |
| EBNA1 |
| Elastin |
| Entaktin EDTA |
| Factor B |
| Factor D |
| Factor H |
| Factor I |
| Factor P |
| Fibrinogen IV |
| Fibrinogen S |
| Fibronectin |
| GAD2/GAD65 |
| GBM |
| Genomic DNA |
| Gliadin |
| Glycated Albumin |
| GP2 |
| GP210 |
| Hemocyanin |
| Heparan sulfate proteoglycan |
| Heparan Sulphate |
| Heparin |
| Histone H1 |
| Histone H2A |
| Histone H2B |
| Histone H3 |
| Histone H4 |
| Insulin |
| Intrinsic Factor |
| Jo-1 |
| KU (P70/P80) |
| La/SSB |
| Laminin |
| LC1 |
| LKM1 |
| LPS |
| M2 |

| |
|---|
| Matrigel |
| MDA5 |
| Mi-2 |
| Mitochondrial antigen |
| MPO |
| Muscarinic receptor |
| Myelin basic protein (MBP) |
| Myosin |
| Nucleolin |
| Nucleosome antigen |
| Nup 62 |
| PCNA |
| Peroxiredoxin 1 |
| PL-7 |
| PL-12 |
| PM/Scl 100 |
| PM/Scl-75 |
| PR3 |
| Proteoglycan |
| Prothrombin protein |
| Ribo Phosphoprotein P0 |
| Ribo Phosphoprotein P1 |
| Ribo Phosphoprotein P2 |
| Ro/SSA (52 Kda) |
| Ro/SSA (60 Kda) |
| S100 |
| Scl-70/Topoisomerase I |
| Sm |
| Sm/RNP |
| SmD |
| SmD1,D2,D3 |
| SP100 |
| Sphingomyelin |
| SRP54 |
| ssDNA |
| ssRNA |
| T1F1 gama |
| Thyroglobulin |
| TNF-a |

| |
|---|
| TPO |
| TTG |
| U1-snRNP 68/70 |
| U1-snRNP A |
| U1-snRNP B/B U1-snRNP B/B |
| U1-snRNP C |
| Vimentin |
| Vitronectin |

| Supplemental Table 3 | | | |
|---|---|---|---|
| **Antibodies** | **Source** | **Catalogue Number** | **Dilutions used** |
| Drp1 (clone 6Z-82) | Santa Cruz | Cat. SC-101270 | 1:100[a]<br>1:1000[b] |
| p53 (clone DO-1) | Santa Cruz | Cat. SC-126 | 1:100 |
| Mfn1 | Santa Cruz | Cat. SC-166644 | 1:100[a]<br>1:1000[b] |
| Mfn2 | Santa Cruz | Cat. SC-515647 | 1:100[a]<br>1:1000[b] |
| PLD6 | Abcam | Cat. ab237612 | 1:100[a]<br>1:1000[b] |
| Miga-1 | Thermo Fischer Scientific | Cat. PA5-53611 | 1:100[a]<br>1:1000[b] |
| LC3-β | Santa Cruz | Cat. SC-376404 | 1:100[a]<br>1:1000[b] |
| Tom-20 | Santa Cruz | Cat. SC-17764 | 1:100[a]<br>1:1000[b] |
| Timm-23 | Santa Cruz | Cat. SC-514463 | 1:100[a]<br>1:1000[b] |
| Mitofilin | Abcam | Cat. ab245764 | 1:100[a]<br>1:1000[b] |
| Myosin IIa | Cell signaling Technology | Cat. 3403 | 1:100[a]<br>1:1000[b] |
| Myosin IIb | Cell signaling Technology | Cat. 3404 | 1:100[a]<br>1:1000[b] |
| EPPK1 | Thermo Fischer Scientific | Cat. PA566913 | 1:100[a]<br>1:1000[b] |
| NF-M (clone E-9) | Santa Cruz | Cat. SC-398532 | 1:1000[b] |
| NF-M (clone 160) | Santa Cruz | Cat. SC-20013 | 1:1000[b] |
| EPLIN (clone 20) | Santa Cruz | Cat. SC-136399 | 1:1000[b] |
| Spectrin A | Santa Cruz | Cat. SC-53444 | 1:1000[b] |

| | | | |
|---|---|---|---|
| Spectrin B | Santa Cruz | Cat. SC-136074 | 1:1000[b] |
| Plectin | Santa Cruz | Cat. SC-33649 | 1:1000[b] |
| Anti-Halo tag | Promega | Cat. 9281 | |
| Anti-Halo tag | Promega | Cat. G912A | |
| Fibronectin-EDA | Santa Cruz | Cat. SC-59826 | 1:1000[b] |
| Fibronectin-P1H11 | Santa Cruz | Cat. SC-18825 | 1:1000[b] |
| Beta Actin (clone 4C2) | Sigma | Cat. MABT825 | 1:100 |
| GAPDH | Santa Cruz | Cat. SC-47724 | 1:100 |
| Anti-Phosphoryl Choline (clone BH8) | Sigma Aldrich | Cat. MABF2084 | |
| Anti-MDA | Sigma Aldrich | Cat. SAB5202544 | |
| Anti-Rabbit Cy-5 secondary antibody | Dianova | Cat. 111-175-144 | 1:100[b] |
| Anti-Mouse Cy-5 secondary antibody | Dianova | Cat. 115-175-146 | 1:100[b] |
| Goat anti-mouse IgG, HRP conjugate | Sigma Aldrich | Cat. 12-349 | 1:10000 |
| Goat anti-rabbit IgG, HRP conjugate | Sigma Aldrich | Cat. 12-348 | 1:10000 |
| Goat anti-human IgM secondary antibody, HRP | Thermo Fischer Scientific | Cat. 31415 | 1:10000 |

[a] dilution for immunofluorescence
[b] dilution for immunoblotting

| Supplemental Table 4 | | |
|---|---|---|
| **Constructs** | **Source (vector backbone)** | **Catalogue number** |
| pLVTHM | addgene | Plasmid #12247 |
| pFN22A-Halo-EBV dUTPase | Promega | NA |
| pFN22A-Halo-HSV1 dUTPase | Promega | NA |
| pFN22A-Halo-HHV6 dUTPase | Promega | NA |
| pFN22A-Halo-Mock dUTPase | Promega | NA |
| pTrcHIS-EBV dUTPase | Thermo Fischer Scientific | NA |
| pTrcHIS-HSV1 dUTPase | Thermo Fischer Scientific | NA |
| pTrcHIS-HHV6 dUTPase | Thermo Fischer Scientific | NA |

**Supplemental Table 5a:** O.D. values at 450 nm for the human IgM standard concentration curves between the range of 0.8 ng/ml to 25.6 ng/ml.

| IgM Concentration (ng/mL) | Assay 1-1 | | | | | Assay 1-2 | | | | |
|---|---|---|---|---|---|---|---|---|---|---|
| | O.D.-1 | O.D.-2 | Mean O.D. | SD | CV | O.D.-1 | O.D.-2 | Mean O.D. | SD | CV |
| 25,600 | 2,998 | 3,010 | 3,004 | 0,0060 | 0,20 | 2,758 | 2,654 | 2,706 | 0,0520 | 1,92 |
| 12,800 | 1,606 | 1,409 | 1,508 | 0,0985 | 6,53 | 1,421 | 1,343 | 1,382 | 0,0390 | 2,82 |
| 6,400 | 0,700 | 0,759 | 0,730 | 0,0295 | 4,04 | 0,621 | 0,656 | 0,639 | 0,0175 | 2,74 |
| 3,200 | 0,322 | 0,378 | 0,350 | 0,0280 | 8,00 | 0,319 | 0,335 | 0,327 | 0,0080 | 2,45 |
| 1,600 | 0,184 | 0,181 | 0,183 | 0,0015 | 0,82 | 0,173 | 0,181 | 0,177 | 0,0040 | 2,26 |
| 0,800 | 0,107 | 0,106 | 0,107 | 0,0005 | 0,47 | 0,108 | 0,125 | 0,117 | 0,0085 | 7,30 |
| 0,000 | 0,044 | 0,045 | 0,045 | 0,0005 | 1,12 | 0,044 | 0,043 | 0,044 | 0,0005 | 1,15 |
| | | | | Mean CV | 3,03 | | | | Mean CV | 2,95 |
| Con. | Assay 2-1 | | | | | Assay 2-2 | | | | |
| | O.D.-1 | O.D.-2 | Mean O.D. | SD | CV | O.D.-1 | O.D.-2 | Mean O.D. | SD | CV |
| 25,600 | 3,025 | 2,783 | 2,904 | 0,1210 | 4,17 | 2,553 | 2,138 | 2,346 | 0,2075 | 8,85 |
| 12,800 | 1,631 | 1,472 | 1,552 | 0,0795 | 5,12 | 1,478 | 1,094 | 1,286 | 0,1920 | 14,93 |
| 6,400 | 0,973 | 0,850 | 0,912 | 0,0615 | 6,75 | 0,584 | 0,585 | 0,585 | 0,0005 | 0,09 |
| 3,200 | 0,450 | 0,464 | 0,457 | 0,0070 | 1,53 | 0,277 | 0,285 | 0,281 | 0,0040 | 1,42 |
| 1,600 | 0,241 | 0,230 | 0,236 | 0,0055 | 2,34 | 0,166 | 0,221 | 0,194 | 0,0275 | 14,21 |
| 0,800 | 0,140 | 0,135 | 0,138 | 0,0025 | 1,82 | 0,105 | 0,097 | 0,101 | 0,0040 | 3,96 |
| 0,000 | 0,044 | 0,045 | 0,045 | 0,0005 | 1,12 | 0,046 | 0,050 | 0,048 | 0,0020 | 4,17 |
| | | | | Mean CV | 3,26 | | | | Mean CV | 6,80 |
| Con. | Assay 3-1 | | | | | Assay 3-2 | | | | |
| | O.D.-1 | O.D.-2 | Mean O.D. | SD | CV | O.D.-1 | O.D.-2 | Mean O.D. | SD | CV |
| 25,600 | 2,480 | 2,137 | 2,309 | 0,1715 | 7,43 | 2,688 | 2,627 | 2,658 | 0,0305 | 1,15 |

| | | | | | | | | | | |
|---|---|---|---|---|---|---|---|---|---|---|
| 12,800 | 1,198 | 1,125 | 1,162 | 0,0365 | 3,14 | 1,430 | 1,352 | 1,391 | 0,0390 | 2,80 |
| 6,400 | 0,568 | 0,578 | 0,573 | 0,0050 | 0,87 | 0,683 | 0,652 | 0,668 | 0,0155 | 2,32 |
| 3,200 | 0,273 | 0,300 | 0,287 | 0,0135 | 4,71 | 0,363 | 0,390 | 0,377 | 0,0135 | 3,59 |
| 1,600 | 0,174 | 0,188 | 0,181 | 0,0070 | 3,87 | 0,196 | 0,195 | 0,196 | 0,0005 | 0,26 |
| 0,800 | 0,097 | 0,097 | 0,097 | 0,0000 | 0,00 | 0,118 | 0,119 | 0,119 | 0,0005 | 0,42 |
| 0,000 | 0,044 | 0,044 | 0,044 | 0,0000 | 0,00 | 0,046 | 0,044 | 0,045 | 0,0010 | 2,22 |
| | | | | Mean CV | 2,86 | | | | Mean CV | 1,82 |
| Con. | Inter-Assay Values | | | | | | | | | |
| | Mean O.D. | SD | CV | | | | | | | |
| 25,600 | 2,654 | 0,2589 | 11,47 | | | | | | | |
| 12,800 | 1,380 | 0,1306 | 5,71 | | | | | | | |
| 6,400 | 0,684 | 0,1143 | 2,86 | | | | | | | |
| 3,200 | 0,346 | 0,0597 | 1,43 | | | | | | | |
| 1,600 | 0,194 | 0,0196 | 0,70 | | | | | | | |
| 0,800 | 0,113 | 0,0134 | 0,34 | | | | | | | |
| 0,000 | 0,045 | 0,0015 | 0,02 | | | | | | | |
| | | Mean CV | 3,22 | | | | | | | |

**Supplemental Table 5b:** O.D. values at 450 nm for the human IgG standard concentration curves between the range of 0.8 ng/ml to 25.6 ng/ml.

| IgG Concentration (ng/mL) | Assay 1-1 | | | | | Assay 1-2 | | | | |
|---|---|---|---|---|---|---|---|---|---|---|
| | O.D.-1 | O.D.-2 | Mean O.D. | SD | CV (%) | O.D.-1 | O.D.-2 | Mean O.D. | SD | CV |
| 25,600 | 2,730 | 2,465 | 2,598 | 0,1325 | 5,10 | 2,213 | 2,342 | 2,278 | 0,0645 | 2,83 |
| 12,800 | 1,390 | 1,209 | 1,300 | 0,0905 | 6,96 | 1,172 | 1,233 | 1,203 | 0,0305 | 2,54 |
| 6,400 | 0,662 | 0,634 | 0,648 | 0,0140 | 2,16 | 0,535 | 0,589 | 0,562 | 0,0270 | 4,80 |
| 3,200 | 0,338 | 0,302 | 0,320 | 0,0180 | 5,63 | 0,266 | 0,256 | 0,261 | 0,0050 | 1,92 |
| 1,600 | 0,176 | 0,181 | 0,179 | 0,0025 | 1,40 | 0,161 | 0,151 | 0,156 | 0,0050 | 3,21 |
| 0,800 | 0,109 | 0,104 | 0,107 | 0,0025 | 2,35 | 0,101 | 0,096 | 0,099 | 0,0025 | 2,54 |
| 0,000 | 0,054 | 0,050 | 0,052 | 0,0020 | 3,85 | 0,048 | 0,048 | 0,048 | 0,0000 | 0,00 |
| | | | | Mean CV | 3,92 | | | | Mean CV | 2,55 |
| Con. | Assay 2-1 | | | | | Assay 2-2 | | | | |
| | O.D.-1 | O.D.-2 | Mean O.D. | SD | CV (%) | O.D.-1 | O.D.-2 | Mean O.D. | SD | CV |
| 25,600 | 2,869 | 2,823 | 2,846 | 0,0230 | 0,81 | 2,257 | 2,297 | 2,277 | 0,0200 | 0,88 |
| 12,800 | 1,372 | 1,325 | 1,349 | 0,0235 | 1,74 | 1,434 | 1,302 | 1,368 | 0,0660 | 4,82 |
| 6,400 | 0,653 | 0,619 | 0,636 | 0,0170 | 2,67 | 0,513 | 0,546 | 0,530 | 0,0165 | 3,12 |
| 3,200 | 0,322 | 0,316 | 0,319 | 0,0030 | 0,94 | 0,292 | 0,260 | 0,276 | 0,0160 | 5,80 |
| 1,600 | 0,182 | 0,177 | 0,180 | 0,0025 | 1,39 | 0,157 | 0,156 | 0,157 | 0,0005 | 0,32 |
| 0,800 | 0,100 | 0,104 | 0,102 | 0,0020 | 1,96 | 0,099 | 0,098 | 0,099 | 0,0005 | 0,51 |
| 0,000 | 0,063 | 0,053 | 0,058 | 0,0050 | 8,62 | 0,052 | 0,050 | 0,051 | 0,0010 | 1,96 |
| | | | | Mean CV | 2,59 | | | | Mean CV | 2,49 |
| Con. | Assay 3-1 | | | | | Assay 3-2 | | | | |
| | O.D.-1 | O.D.-2 | Mean O.D. | SD | CV (%) | O.D.-1 | O.D.-2 | Mean O.D. | SD | CV |
| 25,600 | 3,166 | 2,985 | 3,076 | 0,0905 | 2,94 | 2,819 | 2,702 | 2,761 | 0,0585 | 2,12 |

| | | | | | | | | | | |
|---|---|---|---|---|---|---|---|---|---|---|
| 12,800 | 1,677 | 1,591 | 1,634 | 0,0430 | 2,63 | 1,340 | 1,337 | 1,339 | 0,0015 | 0,11 |
| 6,400 | 0,845 | 0,832 | 0,839 | 0,0065 | 0,78 | 0,617 | 0,597 | 0,607 | 0,0100 | 1,65 |
| 3,200 | 0,441 | 0,440 | 0,441 | 0,0005 | 0,11 | 0,324 | 0,315 | 0,320 | 0,0045 | 1,41 |
| 1,600 | 0,215 | 0,212 | 0,214 | 0,0015 | 0,70 | 0,180 | 0,169 | 0,175 | 0,0055 | 3,15 |
| 0,800 | 0,140 | 0,129 | 0,135 | 0,0055 | 4,09 | 0,106 | 0,107 | 0,107 | 0,0005 | 0,47 |
| 0,000 | 0,053 | 0,047 | 0,050 | 0,0030 | 6,00 | 0,047 | 0,052 | 0,050 | 0,0025 | 5,05 |
| | | | | Mean CV | 2,46 | | | | Mean CV | 1,99 |
| Con. | Inter-Assay Values | | | | | | | | | |
| | Mean O.D. | SD | CV | | | | | | | |
| 25,600 | 2,639 | 0,2920 | 11,48 | | | | | | | |
| 12,800 | 1,365 | 0,1317 | 5,72 | | | | | | | |
| 6,400 | 0,637 | 0,0990 | 2,88 | | | | | | | |
| 3,200 | 0,323 | 0,0576 | 1,44 | | | | | | | |
| 1,600 | 0,176 | 0,0192 | 0,71 | | | | | | | |
| 0,800 | 0,108 | 0,0124 | 0,35 | | | | | | | |
| 0,000 | 0,051 | 0,0032 | 0,03 | | | | | | | |
| | | Mean CV | 3,23 | | | | | | | |
